# Self-applied carrageenan-based gel to prevent human papillomavirus infection in sexually active young women: Final analysis of efficacy and safety of a randomised controlled trial

**DOI:** 10.1101/2023.02.28.23286426

**Authors:** Cassandra Laurie, Mariam El-Zein, Sarah Botting-Provost, Joseph E. Tota, Pierre-Paul Tellier, François Coutlée, Ann N. Burchell, Eduardo L. Franco

## Abstract

**Introduction:** The Carrageenan-gel Against Transmission of Cervical Human papillomavirus trial’s interim analysis (June 2017, n=277) demonstrated a 36% protective effect of carrageenan against incident human papillomavirus (HPV) infections. We report the trial’s final results on efficacy and safety of a carrageenan-based gel in reducing HPV incidence and prevalence.

**Methods:** A phase IIB randomised, placebo-controlled trial, recruited healthy young women aged ≥18 primarily from health service clinics at two Canadian Universities in Montreal, Canada. Participants were randomised (1:1) to a carrageenan-based or placebo gel to be self-applied every other day for the first month and before/after intercourse. Primary outcomes were HPV type-specific incidence and clearance of prevalent infections. At each visit (months 0, 0.5, 1, 3, 6, 9, 12), participants provided questionnaire data and a self-collected vaginal sample (tested for 36 HPV types, Linear Array). Intention-to-treat analyses were conducted using Cox proportional hazards regression models. Incidence and clearance analyses were restricted to participants with ≥2 visits. Trial registration:ISRCTN96104919.

**Findings:** 461 participants (enrolled January 16^th^/2013–September 30^th^/2020) were randomised to carrageenan (n=227) or placebo (n=234) arm. Incidence, clearance, and safety analyses included 429, 240, and 461 participants, respectively. We found 51·9% (108/208) of participants in carrageenan and 66·5% (147/221) in placebo arm acquired ≥1 HPV type (hazard ratio [HR] 0·63 [95% CI: 0·49–0·81]). Among participants who tested HPV-positive at baseline, clearance (two consecutive HPV-negative visits following ≥1 positive visit) was comparable between groups; 31·8% (34/107) in carrageenan and 29·3% (39/133) in placebo arm cleared their infections (HR 1·16 [95% CI: 0·73-1·84]). Adverse events were reported by 34·8% (79/227) and 39·7% (93/234) of participants in carrageenan and placebo arm (p<0.27), respectively.

**Interpretation:** Consistent with the interim analysis, use of a carrageenan-based gel resulted in a 37% reduction in the risk of incident genital HPV infections in women. A carrageenan-based gel may complement HPV vaccination.

**Funding:** Canadian Institute of Health Research (grants MOP-106610 and FDN-143347 to ELF), CarraShield Labs Inc. (St Petersburg, FL) [provided gels in kind].

**RESEARCH IN CONTEXT:** 

**Evidence before this study:** We published in 2021 a narrative review summarizing carrageenan’s preventive effect on human papillomavirus (HPV) based on results from 19 experimental research articles that reported on carrageenan’s anti-HPV activity. Since publication and without applying language or date restriction, we identified four additional records based on a PubMed search using the keywords “carrageenan” and “human papillomavirus” or “HPV” up to January 9th, 2023.

Altogether, these records consist of ten *in vitro* (including 4 *ex vivo*), five *in vitro* and *in vivo*, three *in vivo*, and five clinical studies (including one post-hoc phase III randomised controlled trial [RCT], one observational study, one interim analysis of a phase IIB RCT in women, and two records for the interim analyses of a phase IIB RCT in men). Studies assessed carrageenan alone or in combination with other anti-microbial agents. The results from pre-clinical studies were consistent with a protective effect of carrageenan against HPV.

Overall, eight studies were conducted in humans. The samples collected were either 1) cervicovaginal lavage samples to assess anti-HPV activity *in vitro* (in three *ex vivo* studies, two being phase I RCTs), or 2) genital samples from women (post-hoc RCT [cervical], observational study [sample type not reported], and RCT phase II [vaginal]) or anal samples from men (one study of incidence and one of clearance). In the three *ex vivo* studies, intravaginal use of carrageenan-containing gels was associated with strong anti-HPV activity. In a post-hoc sub study of the trial, a lower HPV prevalence in the carrageenan compared to placebo arm was found at the trial end, but only among the most compliant users (adjusted odds ratio [aOR] 0·62 [95% CI 0·41–0·94], n=348). There were, however, no baseline or intermediate measurements to assess HPV status. An observational study reported that a carrageenan-based gel may accelerate clearance of existing HPV infection (aOR 4·9 [95% CI 1·60–15·1], n=75). The interim analysis of the CATCH study found a 36% protective effect of carrageenan against incident HPV infection(s) (HR 0·64 [95% CI 0·45-0·89], n=277). Conversely, a clinical trial conducted in men who have sex with men did not demonstrate a protective effect of carrageenan on incidence (HR 1·21 [95% CI 0·86–1·70]), or clearance (HR 0·84 [95% CI 0·31–2·27]) of anal HPV infections, and reported more adverse events in the carrageenan (59·8%) relative to the placebo (39·8%) arm.

**Added value of this study:** The CATCH study is the first clinical trial designed to assess the efficacy of a carrageenan-based gel in reducing the risk of incident and prevalent HPV infections in women. Results were consistent when considering HPV subgenera and type-specific analyses. The addition of carrageenan to a lubricant gel does not appear to impact gel tolerability.

**Implications of all the available evidence:** The results of the CATCH trial indicate that carrageenan-based gels could complement HPV vaccination in protecting against HPV-related diseases. Our findings of the clinical efficacy of carrageenan may encourage future research in this area. It would be important to further examine adherence by looking at determinants of adherence, explore the possibility of the addition of carrageenan to condoms, assess the impact of a carrageenan-based gel on anal HPV infections in women, and continue research in the area of multi-purpose prevention technology for agents against HPV, HIV, and other sexually transmitted infections.

## INTRODUCTION

The most common sexually transmitted infection (STI) agent worldwide is human papillomavirus (HPV), which is a necessary cause of cervical cancer.^1^ Globally, cervical cancer is the fourth most commonly diagnosed cancer in women, with an estimated 604,127 new cases and 341,831 deaths in 2020.^2^ Elimination of cervical cancer is believed to be within our reach as per the World Health Organization’s (WHO) 2020 call to eliminate cervical cancer through a three-pronged approach: 1) HPV vaccination of 90% of girls by age 15, 2) cervical cancer screening of 70% of women aged 35–45, and 3) treatment of 90% of women with cervical precancers and cancers.^3^

While HPV vaccination is a highly effective intervention for primary prevention of HPV, the development, implementation, and maintenance of HPV vaccination programs across different settings are not without their challenges, including the negative impact of the COVID-19 pandemic derailing the uptake of HPV vaccines, temporary HPV vaccine shortage, cold chain requirements, and vaccine hesitancy.^4^ Although the global HPV vaccine supply is estimated to meet global demand,^5^ the HPV vaccine shortage highlights the need for the development of complementary primary prevention methods. The use of a personal self-administered gel may support the WHO’s goals to eliminate cervical cancer.

Carrageenan, an anionic polymer derived from red algae, previously showed promise as a potent anti-HPV inhibitor *in vitro* and *in vivo*,^6^ and has a good safety profile for vaginal use.^7^ The Carrageenan-gel Against Transmission of Cervical Human papillomavirus (CATCH) trial was specifically designed to evaluate the efficacy of a carrageenan-based gel against incident and prevalent HPV infections.^8^ Based on the interim analysis (June 2017, N=277), carrageenan-gel use was associated with a 36% protective effect against incident human papillomavirus (HPV) infections compared to placebo gel use.^9^ Anti-microbial agents, such as carrageenan, represent a promising area of research in STI prevention. We now report the results of the final analysis of efficacy and safety of a carrageenan-based gel in reducing HPV incidence and prevalence of genital HPV among sexually-active young women.

## METHODS

The trial findings are reported following the CONSORT (Consolidated Standards of Reporting Trials) 2010 checklist.^10^

### Study design and population

The CATCH trial is an exploratory phase IIB block-randomised, placebo-controlled trial. The full details of the study protocol and data collection were previously described.^13^ We recruited female university students through McGill Health Service Clinic, Concordia Health Services, the Centre intégré de santé et de services sociaux de la Montérégie-Centre-Territoire Champlain-Charles-Le Moyne, and (as of September 2018) at the Gerald Bronfman Department of Oncology at the Division of Cancer Epidemiology of McGill University, Québec, Canada. Enrollment and follow-up visits were conducted at months 0.5, 1, 3, 6, 9, and 12. Study data were securely stored by LFC hosting until December 15, 2019. Thereafter, data were collected and stored using REDCap (Research Electronic Data Capture) tools.^11,12^ The study received ethical approval from McGill University, Centre Hospitalier de l’Université de Montréal, Concordia University, and the Centre intégré de santé et de services sociaux de la Montérégie-Centre-Territoire Champlain-Charles-Le Moyne. Participants gave written informed consent (and e-consent as of September 5, 2019), which was administered by the study nurse.

### Eligibility

Eligible women to participate were those aged 18 years and older, living in Montreal and planning to remain in Montreal for at least the next year, who had vaginal sex with a male partner during the past 3 months and expect do so again in the next 3 months (regardless of whether or not the male partner(s) will change), not currently in a relationship that has lasted longer than 6 months (i.e. likely to be exposed to new HPV infections), willing to follow study instructions and attend follow-up visits for 12 months, who understand French or English, and who use a medically acceptable method of contraception and intend to use it for the duration of the trial. Women were considered ineligible based on the following exclusion criteria: have had a hysterectomy, have a history of cervical lesions/cancer or genital warts, pregnant or planning to immediately become pregnant, currently breast-feeding, had a recent (within the last 6 weeks) pregnancy, abortion, or genital surgery, have human immunodeficiency virus (HIV) infection, have a known allergy or hypersensitivity to vaginal lubricants and have allergy to any of the ingredients of the study product or placebo, have participated in any research studies (past 3 months) related to HPV or cervical cancer, or have participated in any research studies (past 3 months) that require taking medications or supplements, undergo medical tests or procedures, or undertake dietary or exercise regimens.

### Randomisation and masking

Participants were randomised (1:1) to a carrageenan-based or placebo gel by the study nurse, using computer-assisted block randomisation with a block-size of 8. The intervention and placebo gels were packaged in identical containers, except for the product code label; four different codes were used for the intervention gel and an additional four for the control. The participant, study nurses, and laboratory technicians were blinded to group assignment.

### Procedures and data/samples collection

Participants were instructed to apply the study gel every other day for the first month as well as before (and after as of October 26, 2015) each vaginal intercourse. Approximately 5–10mL of the gel was to be applied to the vagina, penis, and/or condom. STI protection methods (e.g., condoms) were encouraged. Participants were asked to refrain from intercourse at least 48 hours prior to their follow-up visits. Both study gels are commercially available (Divine9 and Divine), made by CarraShield Labs Inc. (St Petersburg, FL). The study gels are identical; both are water-based, clear, odourless, tasteless, and of a similar viscosity. The distinguishing feature is the inclusion of carrageenan in the intervention gel.

At enrollment and each follow-up visit, participants completed a computer-assisted questionnaire on sociodemographic characteristics, smoking and alcohol use, medical and sexual history (only at enrollment), sexual activity and study gel use since last visit (only at follow-up), and condom and lubricant use (both enrollment and follow-up). In addition, a daily online calendar included questions on sexual behaviour, gel use, and adverse events (AEs). Participants could enter and modify calendar information for the seven previous days, which required them to log in at least once a week with a username and password. Additional details are provided in Supplementary Section 1: Figure 1 and Tables 1A-1C.

**Figure 1.**
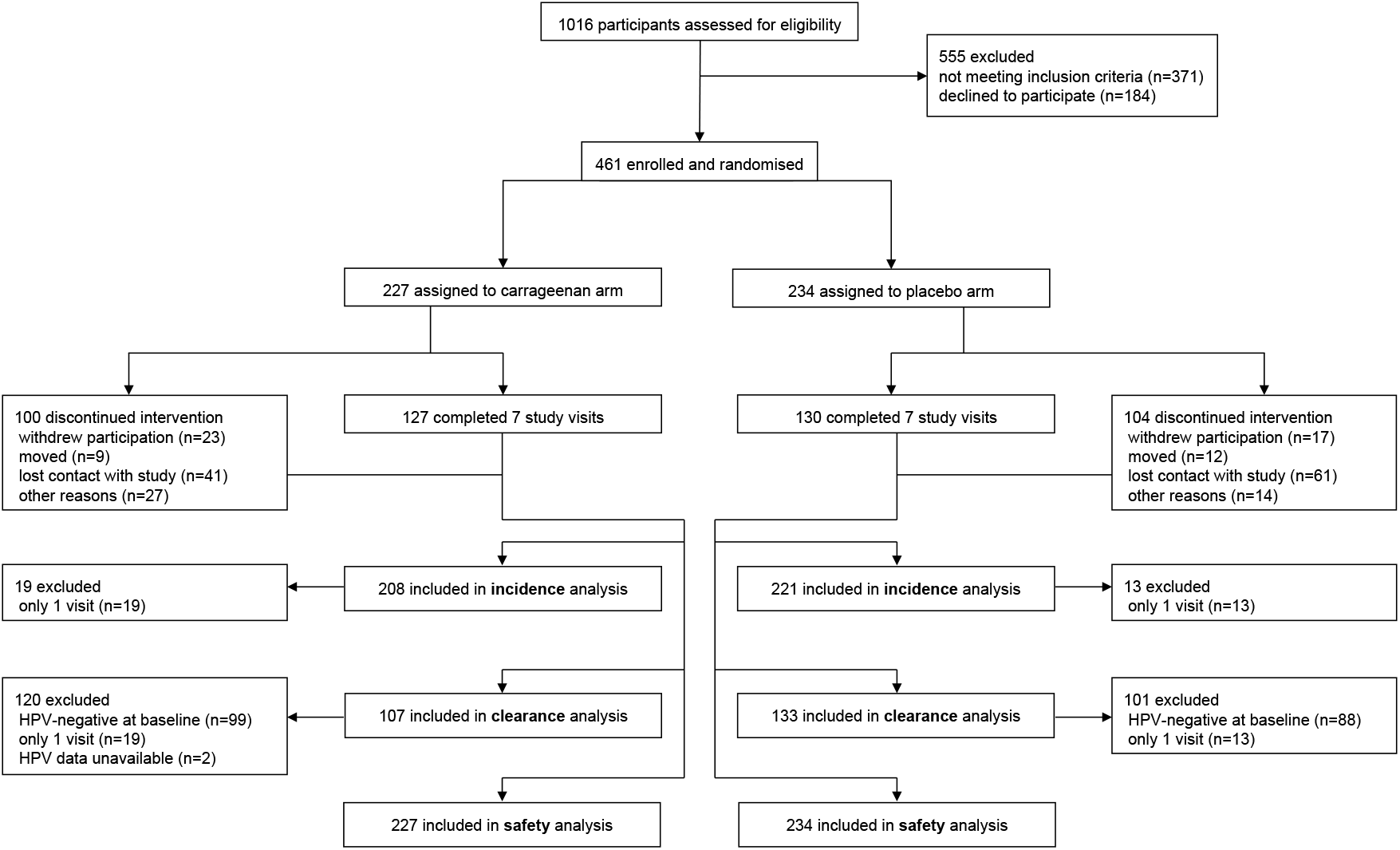
Trial profile: design and subject allocation. The CONSORT flow diagram displays the total number of participants assessed for eligibility who were subsequently enrolled and randomised into either the carrageenan or placebo arm. Reasons for exclusion and discontinuing the intervention are provided. A total of 429 participants were included in the incidence analyses, 240 in the clearance analyses, and 461 in the safety analysis.

**Table 1.** Baseline characteristics of participants, by study arm.

|  | **Carrageenan**<br>**(n=227)** | **Placebo**<br>**(n=234)** |
| --- | --- | --- |
| **Age – years** |  |  |
| Mean (SD) | 25.5 (7.4) | 23.4 (5.4) |
| Median (IQR) | 23.0 (7.4) | 21.9 (5.3) |
| Range | 18.3-68.5 | 18.1-44.2 |
| **Ethnicity, n (%)** |  |  |
| French Canadian | 53 (23.4) | 47 (20.1) |
| English Canadian | 62 (27.3) | 68 (29.1) |
| Black Canadian | 14 (6.2) | 15 (6.4) |
| Latin American | 22 (9.7) | 22 (9.4) |
| South Asian | 9 (4.0) | 7 (3.0) |
| East Asian | 16 (7.1) | 24 (10.3) |
| Other or not reported | 51 (22.5) | 51 (21.8) |
| **Marital status, n (%)** |  |  |
| Single | 145 (63.9) | 160 (68.4) |
| Living with a partner or married | 22 (9.7) | 23 (9.8) |
| Divorced, separated, or widowed | 60 (26.4) | 51 (21.8) |
| **Smoking status, n (%)<sup>a</sup>** |  |  |
| Never | 157 (69.2) | 147 (62.8) |
| Former | 50 (22.0) | 59 (25.2) |
| Current | 19 (8.4) | 27 (11.5) |
| **Age at first intercourse, years** |  |  |
| Mean (SD) | 17.5 (3.0) | 16.9 (2.2) |
| Median (IQR) | 17 (3) | 17 (2) |
| Range | 13-28 | 12-23 |
| Not reported, n | 5 | 5 |
| **Lifetime sex partners – quantiles, n (%)** |  |  |
| <5 | 61 (26.9) | 60 (25.6) |
| 5-7 | 43 (18.9) | 35 (15.0) |
| 8-11 | 37 (16.3) | 47 (20.1) |
| 12-20 | 47 (20.7) | 50 (21.4) |
| ≥ 21 | 39 (17.2) | 42 (18.0) |
| **No. of sex partners in the past month, n (%)<sup>b</sup>** |  |  |
| 0 | 42 (18.5) | 36 (15.5) |
| 1 | 144 (63.4) | 144 (61.8) |
| ≥ 2 | 41 (18.1) | 53 (22.8) |
| **Anal intercourse in the past month, n (%)<sup>c</sup>** |  |  |
| Yes | 25 (11.0) | 26 (11.2) |
| No | 202 (89.0) | 206 (88.8) |
| **HPV DNA status, n (%)** |  |  |
| Any HPV<sup>d</sup> | 115 (50.7) | 141 (60.3) |
| Negative | 109 (48.0) | 93 (39.7) |
| Missing PCR results<sup>e</sup> | 3 (1.32) | 0 (0) |
| Subgenus 1<sup>f</sup> | 38 (17.0) | 55 (23.5) |
| Subgenus 2<sup>g</sup> | 96 (42.9) | 114 (48.7) |
| Subgenus 3<sup>h</sup> | 60 (26.8) | 76 (32.5) |
| **HPV vaccination status, n (%)** |  |  |
| Yes | 97 (42.7) | 120 (51.3) |
| No | 130 (57.3) | 114 (48.7) |
| **Validated HPV vaccination status<sup>i</sup>, n (%)** |  |  |
| Yes | 47 (20.7) | 61 (26.1) |
| No | 64 (28.2) | 63 (26.9) |
| Missing | 116 (51.1) | 110 (47.0) |

Race categories with fewer than 5 participants in each group were collapsed to respect participants anonymity.
<sup>a,b,c</sup> Data were not reported by 2, 1, and 2 participants, respectively.
<sup>d</sup> Participant tested positive for at least 1 of 36 HPV types.
<sup>e</sup> Missing results correspond to invalid or mishandled samples.
<sup>f</sup> Subgenus 1 group includes HPVs 6, 11, 40, 42, 44, and 54.
<sup>g</sup> Subgenus 2 group includes HPVs 16, 18, 26, 31, 33, 34, 35, 39, 45, 51, 52, 53, 56, 58, 59, 66, 67, 68, 69, 70, 73, and 82.
<sup>h</sup> Subgenus 3 group includes HPVs 61, 62, 71, 72, 81, 83, 84, and 89.
<sup>i</sup> Details of the validation of participants' vaccination status can be found in Supplementary section 2 (Figure 2 and Tables 2A-2C).
SD: standard deviation, IQR: interquartile range, PCR: polymerase chain reaction, HPV: human papillomavirus, DNA: deoxyribonucleic acid.

A self-collected vaginal sample was obtained at each study visit. Samples were tested using Linear Array (Roche Molecular Diagnostics, Branchburg, NJ) for detection and genotyping of 36 HPV types.^13^ Samples were tested for beta-globin prior to genotyping. Of 2510 samples, 19 were unavailable (12 samples were beta-globin negative and 7 were mishandled). Carrageenan as used in the clinical protocol above was not shown to inhibit HPV detection in genital samples.^14^

### Outcomes

The two primary outcomes were 1) presence of a newly detected vaginal HPV infection that was not detected at enrollment, and 2) HPV type-specific clearance of infection(s) detected at enrollment. The secondary outcome was adherence (i.e., compliance) to the intervention measured using data from the daily online calendar. Adherent participants were defined as those who used the gel as recommended in >50% of all intercourse acts. While not pre-specified, we also assessed 1) adherence to gel use defined as cumulative adherence prior to censoring or failure, and 2) safety of the study gels by compiling all adverse events that were reported.

### Statistical analysis

Details on sample size calculation in the CATCH protocol showed that the required sample size was 463 for incidence and 388 for clearance analyses.^8^ Based on oncogenic risk and tissue tropism, we grouped HPV types into subgenera; low-risk types in subgenus 1: HPVs 6, 11, 40, 42, 44, and 54; high-risk types in subgenus 2: HPVs 16, 18, 26, 31, 33, 34, 35, 39, 45, 51, 52, 53, 56, 58, 59, 66, 67, 68, 69, 70, 73, and 82; and commensal HPVs in subgenus 3: HPVs 61, 62, 71, 72, 81, 83, 84, and 89.^15-17^ Baseline visits with missing HPV data (n=3) were imputed based on data at the second visit (on average, 14 days after the first visit).

Adherence to the intervention was defined as the number of gel uses divided by the number of vaginal intercourses in the seven days prior to the study visit (from follow-up survey data) and between consecutive study visits (from calendar data). We compared adherence between groups and study visits. Due to (non-significant) imbalances in baseline HPV vaccination status between arms and discrepancies in reporting of vaccination status between screening (pre-enrollment) and the baseline survey (at enrollment), we validated participants’ vaccination status by either confirming their status via email or (in the case of no email reply) by using the date of vaccination reported in their baseline or follow-up survey (Supplementary Section 2: Figure 2 and Tables 2A-2C).

**Figure 2.**
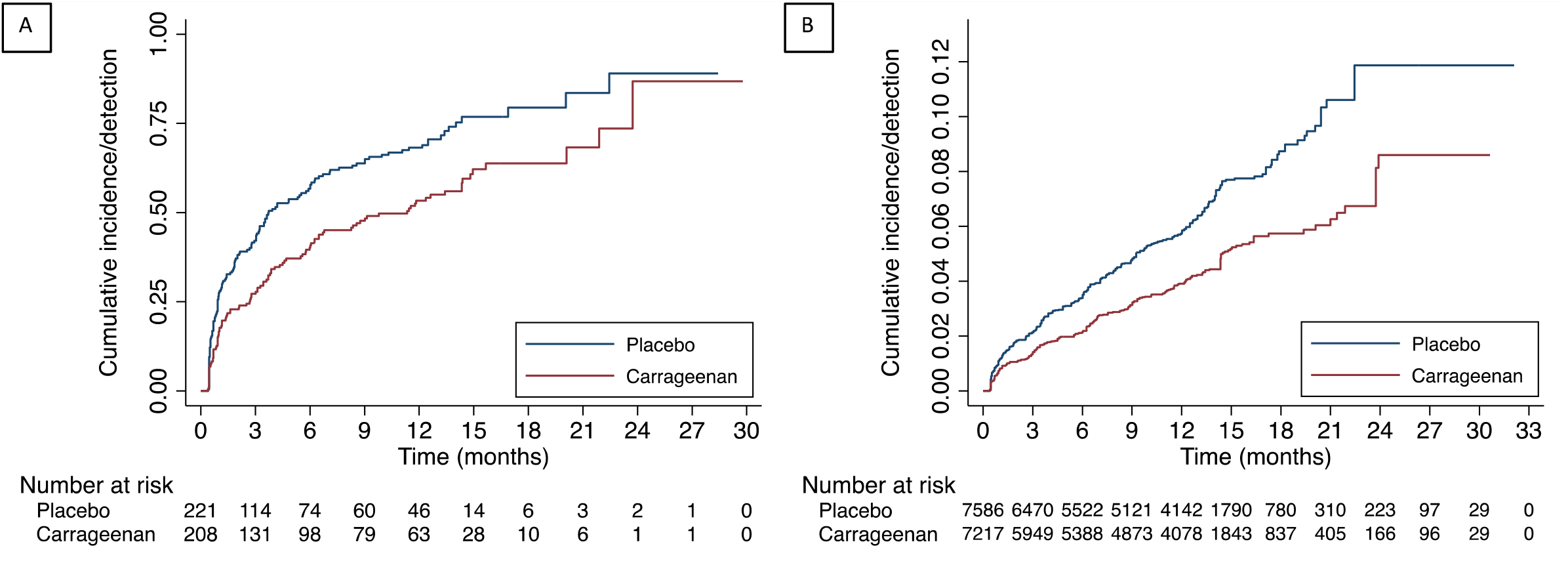
Cumulative incidence/detection of HPV at the participant- and HPV-level. Fig2A plots the first HPV infection episode detected using women as the unit of observation (i.e., participant-level analysis); the number at risk corresponds to the number of women who were HPV-negative for at least 1 of the 36 HPV types at baseline. Fig2B plots all new HPV infections based on HPV-level analysis; the number at risk corresponds to the number of infections a participant could have acquired at subsequent visits. Each participant could have acquired any of the 36 HPV types for which they were negative at baseline.

**Table 2.** Incidence of any HPV infection and grouped infections at the participant- and HPV-level, by study arm.

| Analysis level | HPV infection grouping | Carrageenan |  |  |  | Placebo |  |  |  | Effect estimate |
| --- | --- | --- | --- | --- | --- | --- | --- | --- | --- | --- |
|  |  | N incident/<br>N at risk<br>(%) | Actuarial<br>mean<sup>a</sup><br>(95% CI) | Arithmetic<br>mean<sup>a</sup><br>(95% CI) | Median<sup>a</sup><br>(95% CI) | N Incident/<br>N at risk<br>(%) | Actuarial<br>mean<sup>a</sup><br>(95% CI) | Arithmetic<br>mean<sup>a</sup><br>(95% CI) | Median<sup>a</sup><br>(95% CI) | Hazard ratio<br>(95% CI) |
| **Participant** | Any HPV | 108/208<br>(51.9) | 12.6<sup>b</sup><br>(10.6-14.7) | 4.8<br>(3.8-5.7) | 11.3<br>(6.2-14.3) | 147/221<br>(66.5) | 8.7<sup>b</sup><br>(7.1-10.3) | 3.5<br>(2.8-4.2) | 3.7<br>(3.0-6.0) | 0.63<br>(0.49-0.81) |
|  | Subgenus 1<sup>c</sup> | 33/208<br>(15.9) | 24.8<sup>b</sup><br>(22.9-26.7) | 6.6<br>(4.8-8.3) | NR | 56/221<br>(25.3) | 22.7<sup>b</sup><br>(20.5-24.8) | 5.3<br>(4.0-6.6) | NR<br>(17.8-)<sup>d</sup> | 0.58<br>(0.38-0.89) |
|  | Subgenus 2<sup>e</sup> | 84/208<br>(40.4) | 16.1<sup>b</sup><br>(13.8-18.4) | 5.0<br>(3.8-6.1) | 14.9<br>(12.6-)<sup>d</sup> | 124/221<br>(56.1) | 11.1<br>(9.3-12.9) | 4.4<br>(3.6-5.3) | 8.6<br>(6.0-12.0) | 0.61<br>(0.46-0.81) |
|  | Subgenus 3<sup>f</sup> | 55/208<br>(26.4) | 20.7<sup>b</sup><br>(18.3-23.1) | 5.7<br>(4.2-7.3) | 23.7<br>(20.1-)<sup>d</sup> | 79/221<br>(35.8) | 19.2<sup>b</sup><br>(16.8-21.6) | 5.5<br>(4.4-6.5) | 22.4<br>(13.6-)<sup>d</sup> | 0.70<br>(0.50-0.99) |
| **HPV** | Any HPV<sup>g</sup> | 278/7,217<br>(3.9) | 29.1<sup>b</sup><br>(28.9-29.3) | 6.7<br>(6.0-7.3) | NR | 438/7,586<br>(5.8) | 29.7<sup>b</sup><br>(29.4-29.9) | 6.4<br>(5.9-6.9) | NR | 0.65<br>(0.50-0.83) |

<sup>a</sup> Time in months. The actuarial mean accounts for censoring, whereas the arithmetic mean excludes participants who did not acquire a new HPV type.
<sup>b</sup> Mean was underestimated since the largest observed analysis time was censored.
<sup>c</sup> Subgenus 1 includes HPV types 6, 11, 40, 42, 44, and 54.
<sup>d</sup> Upper confidence limit was undetermined since the survival function did not fall below 0.5.
<sup>e</sup> Subgenus 2 includes HPV types 16, 18, 26, 31, 33, 34, 35, 39, 45, 51, 52, 53, 56, 58, 59, 66, 67, 68, 69, 70, 73, and 82.
<sup>g</sup> Proportional hazards Cox regression models account for all incident HPV infections acquired over follow-up. Participants were considered at risk for any HPV type absent at baseline. Each participant could contribute up to 36 observations, each corresponding to an HPV type. The unit of analysis was each individual HPV type.
CI: confidence interval, HPV: human papillomavirus, NR: not reached, N: number

Cox proportional hazards regression models were used to evaluate the efficacy of carrageenan in reducing vaginal HPV incidence/detection and prevalence. Missing HPV data for follow-up visits were censored in incidence and clearance analyses. Incidence analyses, performed by any HPV positivity (if a participant was positive for one or more HPV types at baseline, they remain at risk of acquiring HPV types absent at baseline) and subgenera (separate analyses done for each of the three subgenera), included participants who had at least 1 follow-up visit. We performed sub-group analyses to compare the intervention effects according to main baseline characteristics and cumulative adherence to gel use prior to failure (new detection of an HPV type) or censoring.

Participants were included in clearance analyses if they had at least two study visits and were HPV-positive at baseline. We considered two definitions of clearance: liberal (one HPV-negative visit following at least one HPV-positive visit) and conservative (two consecutive HPV-negative visits following at least one HPV-positive visit). We considered time to 1) clearance of all baseline HPV infections and 2) first clearance of any baseline HPV infection.

Analyses were also done at the HPV-level, where the unit of analysis was each individual HPV type. The models were stratified by HPV type and clustered by participant. For incidence analyses, participants were considered at risk for any HPV type absent at baseline, where each participant could contribute up to 36 observations, each corresponding to an HPV type. For HPV-level clearance analyses, participants were considered at risk for clearing any of their baseline HPV infections.

To assess safety of the study gels, we tabulated self-reported AEs and/or reactions that were recorded by means of the 1) *daily calendar*, 2) *follow-up survey* administered at each follow-up visit, 3) *adverse event module (nurse report)* during follow-up visits, and/or 4) reporting of an adverse event during *adverse event follow-up*. AEs reported in the daily calendar were graded (mild, moderate, or severe) by participants.

Data management and analyses were done in SAS V9.4 and Stata version 17, respectively. The trial was registered (ISRCTN96104919) and approved by Health Canada (authorization file number 169160). A data safety and monitoring board reviewed and recommended publication of the previously published interim analysis,^9^ as well as the final trial results reported herein.

## RESULTS

Recruitment took place between January 16th, 2013 and October 30th, 2020. As shown in Figure 1, 1016 participants were assessed for eligibility. Of these, 461 were randomised into the carrageenan (n=227) and placebo (n=234) arms. Study visits 2–7 were completed by 461, 429, 399, 362, 318, 291, and 257 participants, respectively. Overall, 429, 240, and 461 participants were included in the incidence, clearance, and safety analyses, respectively. Supplementary Figure 3 shows the distribution of time between baseline and subsequent visits compared to the study schedule. While there were variations between the actual and planned study schedule, little difference was observed between intervention arms.

Baseline characteristics of study participants are shown in Table 1. The median age was 23·0 in the carrageenan and 21·9 in the placebo arm. Most participants were Canadian, single, never smokers, had less than 5 lifetime sexual partners, and had 1 sexual partner in the past month. The median age at first intercourse was 17. More participants in placebo (60·3%) compared to carrageenan (50·7%) arm were infected with any HPV type at baseline, which was consistent by subgenera. Detailed HPV prevalence data at each study visit are shown for carrageenan and placebo arms in Supplementary Tables 3A and B, respectively. Fewer participants in the carrageenan (26·1%) compared to placebo arm (31·7%) reported ever having anal intercourse during follow-up. Fewer participants in carrageenan (42·7%) compared to placebo (51·3%) arm reported being vaccinated against HPV. The difference in vaccination status between arms was smaller based on validated vaccination status; 42·3% of participants in carrageenan and 49·2% in placebo arm reported receiving the HPV vaccine.

Follow-up characteristics of the participants are shown in Supplementary Table 4. The median overall follow-up time was 12·2 months. At the observation level (observations between two consecutive study visits), adherence was comparable between arms; 26·0% of participants in carrageenan and 26·9% in placebo arm reported >75% adherence in the 7 days preceding each visit based on data from the follow-up surveys.

As shown in Table 2, fewer participants in carrageenan (51·9%, 108/208) acquired an incident HPV infection compared to placebo (66·5%, 147/221) arm (HR 0·63, 95% CI 0·49–0·81). This effect was consistently observed in participant-level analyses by subgenera (HRs ranged from 0·58–0·70) and in the HPV-level analysis (HR 0·65, 95% CI 0·50–0·83). Figure 2 shows the cumulative incidence of HPV by intervention arm; participant- (Fig2A) and HPV- (Fig2B) level results were consistent with a protective effect of carrageenan against incident HPV infections.

Table 3 shows time to clearance of all HPV types and time to clearance of the first cleared HPV infection by clearance definition (liberal and conservative). The effect estimates were consistently greater than the null value but never reached statistical significance, irrespective of the analysis level, clearance outcome, or clearance definition. Based on liberal clearance definition of all HPV types, 57·9% (62/107) of participants cleared all their baseline infections in carrageenan and 49·6% (66/133) in placebo arm, corresponding to a HR of 1·41 (95% CI 0·99–2·00). By applying the conservative definition, the HR was 1·16 (95% CI 0·73–1·84). When considering only the first cleared infection, 79·4% (85/107) cleared at least 1 HPV infection in the carrageenan and 79·0% (105/133) in the placebo arm (HR 1·03, 95% CI 0·77–1·38) for the liberal definition; these values were 62·6% (67/107) and 60·9% (81/133) (HR 1·04, 95% CI 0·75–1·44) for conservative clearance, respectively. No remarkable differences were observed when considering time to clearance of individual HPV types; 66·8% cleared at least 1 HPV type in the carrageenan and 65·7% in the placebo arm (HR 1·17, 95% CI 0·90–1·51) for the liberal definition, and 45·0% cleared at least 1 HPV type in the carrageenan and 45·7% in the placebo arm (HR 1·10, 95% CI 0·84–1·43) for the conservative definition.

**Table 3.** Clearance, according to outcome and definition, of any HPV infection at the participant- and HPV-level, by study arm.

| Analysis level | Outcome | Clearance definition<sup>a</sup> | Carrageenan |  |  |  | Placebo |  |  |  | Effect estimate |
| --- | --- | --- | --- | --- | --- | --- | --- | --- | --- | --- | --- |
|  |  |  | N cleared/<br>N at risk<br>(%) | Actuarial<br>mean<sup>b</sup><br>(95% CI) | Arithmetic<br>mean<sup>b</sup><br>(95% CI) | Median<sup>b</sup><br>(95% CI) | N cleared/<br>N at risk<br>(%) | Actuarial<br>mean<sup>b</sup><br>(95% CI) | Arithmetic<br>mean<sup>b</sup><br>(95% CI) | Median<sup>b</sup><br>(95% CI) | Hazard ratio<br>(95% CI) |
| Participant | Time to clearance of all HPV types<sup>c</sup> | Liberal | 62/107<br>(57.9) | 10.0<br>(8.4-11.6) | 5.4<br>(4.8-5.9) | 9.3<br>(6.4-11.0) | 66/133<br>(49.6) | 13.0<br>(10.9-15.0) | 5.8<br>(5.2-6.4) | 11.1<br>(9.5-13.4) | 1.41<br>(0.99-2.00) |
|  |  | Conservative | 34/107<br>(31.8) | 14.8<sup>d</sup><br>(13.0-16.6) | 5.7<br>(5.0-6.5) | 16.0<br>(10.5-NR) | 39/133<br>(29.3) | 19.3<br>(16.8-21.7) | 6.2<br>(5.4-7.0) | 28.9<br>(13.6-NR) | 1.16<br>(0.73-1.84) |
|  | Time to first cleared HPV infection<sup>e</sup> | Liberal | 85/107<br>(79.4) | 4.7<sup>d</sup><br>(3.6-5.8) | 5.0<br>(4.6-5.5) | 3.0<br>(1.2-4.1) | 105/133<br>(79.0) | 4.8<br>(3.9-5.7) | 5.4<br>(4.9-5.8) | 3.2<br>(1.6-4.0) | 1.03<br>(0.77-1.38) |
|  |  | Conservative | 67/107<br>(62.6) | 6.8<sup>d</sup><br>(5.4-8.1) | 5.4<br>(4.9-6.0) | 6.0<br>(4.1-6.5) | 81/133<br>(60.9) | 7.6<sup>d</sup><br>(6.1-9.2) | 5.7<br>(5.2-6.2) | 5.8<br>(3.2-6.9) | 1.04<br>(0.75-1.44) |
| HPV | Time to clearance of an individual HPV type<sup>f</sup> | Liberal | 181/271<br>(66.8) | 0.24<br>(0.2-0.3) | 5.0<br>(4.2-5.7) | 0.20<br>(0.1-0.2) | 243/370<br>(65.7) | 0.29<br>(0.26-0.32) | 5.9<br>(5.3-6.5) | 0.23<br>(0.21-0.25) | 1.17<br>(0.90-1.51) |
|  |  | Conservative | 122/271<br>(45.0) | 0.38<sup>d</sup><br>(0.3-0.4) | 5.1<br>(4.4-5.9) | 0.31<br>(0.3-0.4) | 169/370<br>(45.7) | 0.45<br>(0.40-0.50) | 5.6<br>(4.8-6.3) | 0.34<br>(0.27-0.38) | 1.10<br>(0.84-1.43) |

<sup>a</sup> Liberal clearance was defined as having a single HPV-negative visit following $\geq 1$ HPV-positive visit(s). Conservative clearance was defined as having $\geq 2$ consecutive HPV-negative visits following $\geq 1$ HPV-positive visit(s).
<sup>b</sup> Time in months. The actuarial mean accounts for censoring, whereas the arithmetic mean excludes participants who did not acquire a new HPV type.
<sup>c</sup> Time to clearance of all baseline HPV infections (i.e., clearance was considered to have occurred once all baseline HPV infections cleared).
<sup>d</sup> Mean was underestimated since the largest observed analysis time was censored.
<sup>e</sup> Time to clearance of the first baseline HPV infection (i.e., clearance was considered to have occurred once the first of any baseline HPV infections cleared).
<sup>f</sup> Proportional hazards Cox regression models account for all baseline HPV types that cleared over follow-up. Participants were considered at risk for clearing any HPV type present at baseline. Each participant could contribute up to 36 observations, each corresponding to an HPV type. The unit of analysis was each individual HPV type.
CI: confidence interval, HPV: human papillomavirus, NR: not reached, N: number.

Overall, there was a similar proportion of AEs reported in each arm: 34·8% (79/227) in the carrageenan and 39·7% in the placebo arm (93/234) (Table 4). Those most frequently reported were vaginal yeast infection, itching, burning or pain in the genital area, and bacterial vaginosis.

**Table 4.**
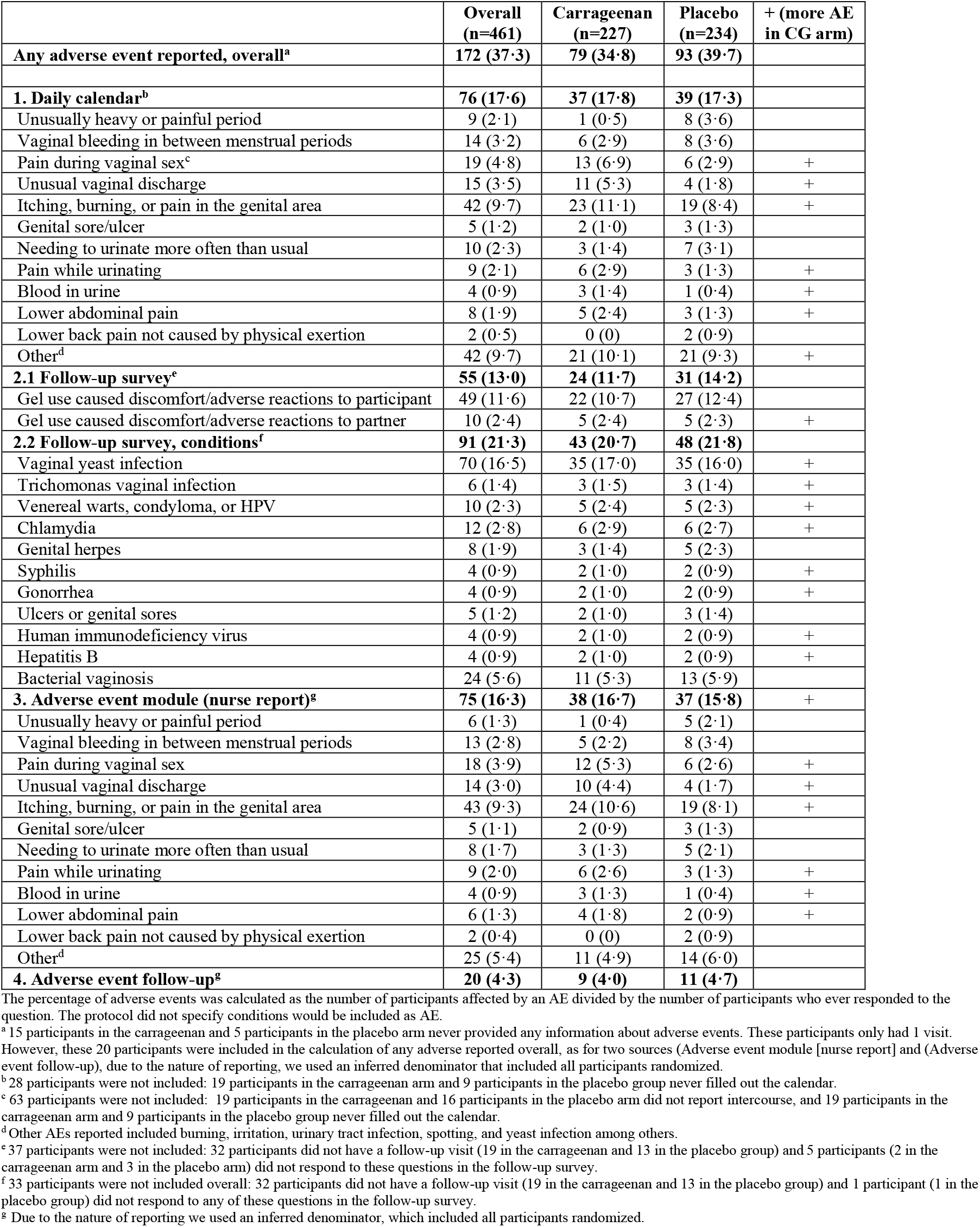
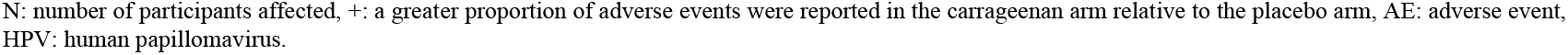
Adverse events [n (%)] reported through different sources, overall and by study arm.

Of participants who reported an AE in the daily calendar, 24·3% (9/37) in carrageenan and 30·8% (12/39) in placebo arm graded their AEs as severe (Supplementary Table 5); the most common severe AE was “itching, burning or pain in the genital area”. We did not observe a higher proportion of withdrawals among participants who reported adverse events nor among participants who reported difficulties using the study gel (Supplementary Table 6). The proportion of participants reporting difficulties with the study gel before, during, or after intercourse was similar between arms (77·2% [159/204] in the carrageenan and 80·7% [176/218] in the placebo arm); the most frequently reported difficulties were not having the CATCH gel at the time of intercourse or forgetting to use the CATCH gel (Supplementary Table 7).

No differences were observed in sub-group analyses by baseline characteristics (age, ethnicity, marital status, smoking status, age at first intercourse, lifetime sexual partners, sex partners in the last month, HPV status at baseline, and vaccination status) and cumulative adherence to gel use (Supplementary Figure 4). There was also no evidence of a dose-response relationship based on cumulative adherence to gel use prior to failure or censoring, irrespective of data used (calendar data or follow-up survey data) nor categorization (binary [<50, ≥50] or categorical), nor was a more protective effect observed when analyses were restricted to participants with an overall adherence >50% (per-protocol analysis) [Supplementary Table 8].

## DISCUSSION

We found a 37% protective effect of carrageenan-gel use against incident HPV infections compared to placebo gel use, but no acceleration of clearance of existing HPV infections. Adherence was balanced between arms, and the study gels were generally well tolerated. These findings corroborate the interim analysis results, published in 2019, which reported a nearly identical estimate of a 36% protective effect of carrageenan against incident HPV infection (HR 0·64 [95% CI 0·45-0·89], N=277).^9^

Clinical studies, and trials in particular, on the efficacy of carrageenan-based gels against HPV infection are scarce (Supplementary Table 9). The CATCH study represents the first phase IIB trial specifically designed to assess the efficacy of carrageenan against incident and prevalent HPV infections. Prior to its initiation, a post-hoc analysis of a trial originally designed to assess carrageenan’s anti-HIV effect found a lower prevalence of HPV at the trial’s end in compliant users of Carraguard® (carrageenan gel) compared to compliant placebo (methylcellulose) gel users (aOR 0·62 [95% CI 0·41–0·94], N=348) adjusted for site, STI, average coital frequency, longer time in study, abnormal pap smear, baseline condom use, age by relationship, and promiscuity by condom use; however, this was a subgroup analysis, and there were no baseline nor intermediate measures of HPV to use as a reference.^18^ Other smaller trials, evaluated carrageenan’s anti-HPV activity *in vitro* from a cervicovaginal lavage using combination products of MIV-150, zinc acetate dihydrate, and carrageenan (20 participants),^19^ as well as carrageenan and griffithsin (13 participants),^20^ demonstrating anti-HPV activity in all or the majority of samples. Moreover, while there was evidence for carrageenan preventing vaginal HPV infections, carrageenan-based gels have not been shown to be effective against anal HPV infections in men,^21^ possibly due in part to anatomical differences between the vagina and anal canal, as well as histological differences: stratified squamous in the vagina and simple columnar and stratified squamous epithelium in the anal canal.

Adherence to the intervention was balanced by arm. We would expect to see that more compliant participants would have increased protection against incident HPV infections. Another trial, assessing Carraguard®’s anti-HPV activity post-hoc, found a protective effect of carrageenan against prevalent HPV infections but only among adherent users (20·3%, 348/1718).^18^ However, in the CATCH trial, a dose-response relationship was not observed. This could be due to misclassification of adherence. For example, during the first month of participation, participants were asked to apply the gel every other day for the purpose of assessing if the gel could increase clearance of existing infections. If a participant reported vaginal intercourse but no gel use during vaginal intercourse, but used the gel outside of vaginal intercourse, she would be classified as non-compliant but may have been offered some protection against HPV infection due to residual carrageenan gel in the vaginal canal. One study showed that carrageenan is present 8–24 hours after Carraguard® gel application for the majority of participants (75%, 12/16), based on cervicovaginal lavage samples.^22^ In a study that tested Divine9™, carrageenan was detected in mouse vaginal washes up to 4 hours after application, but not after 24 hours.^23^ Another study testing Divine9, but in cervicovaginal lavage samples, detected carrageenan in the majority (78%) of samples 8 hours after gel application.^24^ Based on these studies, it is reasonable to expect that there would at least be some anti-HPV activity several hours post-application, suggesting that if a woman applied the gel in the morning and had intercourse in the evening that she could have been offered some protection.

The assessment of gel safety, comparing Divine9 (intervention) to Divine (placebo), found 37% of participants reported AEs. A large trial (N=6202), comparing Carraguard® (a carrageenan-based gel) to methylcellulose with up to 2 years of follow-up, reported that 23% of participants experienced AEs.^7^ This indicates worse tolerability of the study gels used in the CATCH trial compared to the Carraguard® trial. Over the course of follow-up, 16·5% of CATCH participants reported a vaginal yeast infection and 5·6% reported having bacterial vaginosis. These proportions are lower than those reported in a phase II safety trial (N=165), comparing Carraguard® to methylcellulose, where 38% of participants reported having a yeast infection and 22% reported having bacterial vaginosis over follow-up.^25^ The difference may be explained by the nature of reporting; these conditions were self-reported in the CATCH study, whereas they were systematically tested for at each study visit in the safety trial.^25^ The nature of reporting may have led to detection bias (i.e., systematically testing for these conditions leading to more detections), as bacterial vaginosis can be asymptomatic and therefore not self-reported by participants.

While the study gels were approved by Health Canada and have received 510(k) clearance by the United Stated Food and Drug Administration (FDA), they are hyperosmolal and therefore do not meet the current WHO recommendation for osmolality (<1200mOsM/kg).^26^ Hyperosmolality and certain ingredients, such as polyquaternium-15 and nonoxynol-9, have been shown to increase epithelial exfoliation, or sloughing, and cause cytotoxicity, potentially increasing the risk of acquiring certain STIs.^26^ However, these products remain commercially available. Studying the tolerability of lubricants and identifying the ingredients responsible for causing irritation is a challenge, as conclusive evidence on the safety and harms associated with personal lubricant is difficult to generate due to a high number of potential confounders, such as condom use, frequency of sex, and differences in reporting of lubricant use. For example, a lack of evidence of harm caused by polyquaternium led the WHO in 2020 to revert its 2012 recommendation to avoid polyquaternium in personal lubricants.^26^ The study gels were later reformulated using the same type of carrageenan at the same concentration but with adjustments to inactive ingredients to meet the WHO recommendations on osmolality. The reformulated gel was not used in the CATCH trial (Dean Fresonke, Personal communication).

The trial was not without limitations. First, gel use was self-reported, which may be subject to social desirability bias if participants tended to overreport adherence to gel use. This could possibly lead to misclassification of exposure and would be expected to bias the estimate towards the null in adherence analyses. Second, the proportion of AEs experienced by participants in the carrageenan arm might have been underestimated. Comparing participants in carrageenan to placebo arm, there were fewer participants randomised (227 versus 234), more participants who discontinued participation after their first visit (19/227 versus 13/234), and fewer participants who reported AEs (19/227 versus 9/234), leading to fewer participants available to report AEs in the carrageenan arm. Thirdly, the placebo gel, while identical to the intervention gel except for the addition of carrageenan, may not be a true placebo, as it could have anti-HPV activity that would bias the estimate towards the null. While AEs were balanced between arms, this does not exclude the possibility that the study gels could have caused more irritation than would be expected from other common lubricants or methylcellulose, which has been used in other microbicide trials. The greatest strength of the trial is that it is the first phase IIB randomized controlled trial designed to assess the efficacy of a carrageenan-based gel on incident and prevalent HPV infections. A strength of having two identical gels, with the exception of the addition of carrageenan, is that the protective effect observed can be attributed to the presence of carrageenan. An additional strength is the collection of detailed information over time, in particular the implementation of a daily calendar to obtain details of gel use, sexual activity, and adverse events, which lead to the collection of 95,327 observations. HPV positivity was assessed by detecting 36 different HPV types using the Linear Array, which allowed us to assess genotype-specific HPV incidence among HPV-positive women.

It has been hypothesized that carrageenan works by binding directly to the HPV capsid, thereby preventing HPV from binding to heparan sulfate proteoglycan (HSPG), which is a key step in the infection process on the basement membrane.^27^ Carrageenan has also been shown to prevent attachment of HPV to human sperm,^28^ potentially preventing dispersion of HPV within the vaginal canal. Carrageenan may also act through a secondary mechanism independent of HSPG, where carrageenan prevents the virus from binding secondary receptors during the infection process.^28,29^ Multiple mechanisms may be responsible for the anti-HPV activity demonstrated in the CATCH study.

This research supports current investigations on the use and utility of carrageenan co-formulated with other anti-microbials (such as griffithsin and carrageenan) in multi-prevention technologies,^20^ as well as with MIV-150, consisting of an anti-HIV agent, zinc acetate and carrageenan.^19^ To improve adherence to the intervention and encourage STI protection in general, it may be equally important to also study use of carrageenan-based gels in pre-packaged condoms.

Despite the high osmolality of the study gels, carrageenan was protective against incident HPV infections in women. Given that carrageenan-based gels have not demonstrated efficacy in preventing anal HPV infections in men, it would be necessary to conduct a gel tolerability study with the reformulated carrageenan-based gel at the correct osmolality in both women and men that include measurements of anal HPV.

In conclusion, carrageenan compared to placebo gel use was associated with a reduction in incident HPV infections. Topical microbicides against HPV, such as carrageenan-based gels, may have utility as a complement to HPV vaccination among sexually active women.

## Supporting information

CATCH Supplement

CATCH Consort Checklist

## Data Availability

All data produced in the present study will be made available online at the time of submission for publication and can be found in the McGill Dataverse at

https://doi.org/10.5683/SP3/0DS6FP

## Contributors

The authors contributed to: conceptualisation (ELF, ANB, FC, P-PT, JET, CL, MZ); data curation (MZ, CL, SBP); formal analysis (CL); funding acquisition (ELF, ANB, FC, P-PT, JET); investigation (ELF, MZ, CL, SBP, FC), methodology (ELF, ANB, FC, P-PT, JET, MZ, CL); project administration (ELF, MZ); resources (ELF, MZ, FC); supervision (ELF, MZ); validation (CL, SBP); visualisation (CL); writing – original draft (CL); and writing – reviewing and editing (ELF, ANB, FC, P-PT, JET, MZ, SBP, CL). CL and SBP directly assessed and verified the underlying data. All authors had full access to the study data and had the final responsibility to decide to submit for publication.

## Data sharing

The sample code used to generate the results are available in the McGill Dataverse repository (https://doi.org/10.5683/SP3/0DS6FP). Following publication, the anonymized study data and a codebook will be made also available. The study protocol and accompanying documents, such as the informed consent form were previously published.^8^

## Acknowledgements

The study was funded by the Canadian Institutes of Health Research (grant number MOP-106610 and grant FDN-143347 to ELF). CarraShield Labs Inc. (St Petersburg, FL) provided the gels in-kind. CarraShield Labs Inc was not involved in the design of the study, data collection, analyses, and interpretation of trial results. ANB is a Canada Research Chair in Sexually Transmitted Infection Prevention; she also receives salary support from a Non-Clinician Scientist Award, Department of Family and Community Medicine, Faculty of Medicine, University of Toronto.

We wish to thank study participants, employees of the CATCH study, and laboratory research assistants. We also acknowledge the members of the Data Safety and Monitoring Board who evaluated the safety and efficacy of the CATCH study at the interim and final analysis stages.

## REFERENCES

1. de Sanjosé S, Diaz M, Castellsagué X, et al. Worldwide prevalence and genotype distribution of cervical human papillomavirus DNA in women with normal cytology: a meta-analysis. Lancet Infect Dis 2007; 7: 453–9.

2. Sung H, Ferlay J, Siegel RL, et al. Global Cancer Statistics 2020: GLOBOCAN Estimates of Incidence and Mortality Worldwide for 36 Cancers in 185 Countries. CA Cancer J Clin 2021; 71(3): 209–49.

3. WHO. A cervical cancer-free future: First-ever global commitment to eliminate a cancer. 2020. https://www.who.int/news/item/17-11-2020-a-cervical-cancer-free-future-first-ever-global-commitment-to-eliminate-a-cancer#:~:text=eliminate%20a%20cancer-,A%20cervical%20cancer%2Dfree%20future%3A%20First%2Dever%20global,commitment%20to%20eliminate%20a%20cancer&text=WHO's%20Global%20Strategy%20to,%3A%20vaccination%2C%20screening%20and%20treatment. (accessed December 26, 2022).

4. Toh ZQ, Russell FM, Garland SM, Mulholland EK, Patton G, Licciardi PV. Human Papillomavirus Vaccination after COVID-19. JNCI Cancer Spectr 2021; 5.

5. WHO. Global Market Study HPV (Working Document) March 2022. 2022. https://cdn.who.int/media/docs/default-source/immunization/mi4a/who-mi4a-global-market-study-hpv.pdf?sfvrsn=649561b3_1&download=true (accessed December 26, 2022).

6. Laurie C, El-Zein M, Coutlée F, de Pokomandy A, Franco EL. Carrageenan as a preventive agent against human papillomavirus infection. Sex Transm Dis 2021; 48: 458–65.

7. Skoler-Karpoff S, Ramjee G, Ahmed K, et al. Efficacy of Carraguard for prevention of HIV infection in women in South Africa: a randomised, double-blind, placebo-controlled trial. Lancet 2008; 372: 1977–87.

8. Laurie C, Tota JE, El-Zein M, et al. Design and methods for the Carrageenan-gel Against Transmission of Cervical Human papillomavirus (CATCH) study: A randomized controlled trial. Contemp Clin Trials 2021; 110: 106560.

9. Magnan S, Tota JE, El-Zein M, et al. Efficacy of a Carrageenan gel Against Transmission of Cervical HPV (CATCH): interim analysis of a randomized, double-blind, placebo-controlled, phase 2B trial. Clinic Microbiol Infect 2019; 25: 210–6.

10. Schulz KF, Altman DG, Moher D, Group C. CONSORT 2010 Statement: Updated Guidelines for Reporting Parallel Group Randomized Trials. Ann Intern Med 2010; 152(11): 726–32.

11. Harris PA, Taylor R, Thielke R, Payne J, Gonzalez N, Conde JG. Research electronic data capture (REDCap)—A metadata-driven methodology and workflow process for providing translational research informatics support. J Biomed Inform 2009; 42: 377–81.

12. Harris PA, Taylor R, Minor BL, et al. The REDCap consortium: Building an international community of software platform partners. J Biomed Inform 2019; 95: 103208.

13. Coutlée F, Rouleau D, Petignat P, et al. Enhanced detection and typing of human papillomavirus (HPV) DNA in anogenital samples with PGMY primers and the linear array HPV genotyping test. J Clin Microbiol 2006; 44: 1998–2006.

14. Laurie C, El-Zein M, Franco EL, Coutlée F. Assessment of the possible inhibitory effect of carrageenan in human papillomavirus DNA testing by polymerase chain reaction amplification. J Med Virol 2021; 93: 6408–11.

15. Shaw E, Ramanakumar AV, El-Zein M, et al. Reproductive and genital health and risk of cervical human papillomavirus infection: Results from the Ludwig-McGill cohort study. BMC Infect Dis 2016; 16: 116.

16. Schiffman M, Clifford G, Buonaguro FM. Classification of weakly carcinogenic human papillomavirus types: addressing the limits of epidemiology at the borderline. Infect Agent Cancer 2009; 4: 8.

17. Schiffman M, Herrero R, DeSalle R, et al. The carcinogenicity of human papillomavirus types reflects viral evolution. Virology 2005; 337: 76–84.

18. Marais D, Gawarecki D, Allan B, et al. The effectiveness of Carraguard, a vaginal microbicide, in protecting women against high-risk human papillomavirus infection. Antivir Ther 2011; 16: 1219–26.

19. Friedland BA, Hoesley CJ, Plagianos M, et al. First-in-Human Trial of MIV-150 and Zinc Acetate Coformulated in a Carrageenan Gel. J Acquir Immune Defic Syndr 2016; 73: 489– 96.

20. Teleshova N, Keller MJ, Romero JAF, et al. Results of a phase 1, randomized, placebocontrolled first-in-human trial of griffithsin formulated in a carrageenan vaginal gel. PLoS One 2022; 17: e0261775.

21. Laurie C, El-Zein M, Tota JE, et al. Efficacy of a carrageenan gel in preventing anal human papillomavirus (HPV) infection: interim analysis of the Lubricant Investigation in Men to Inhibit Transmission of HPV Infection (LIMIT-HPV) randomised controlled trial. Sex Transm Infect 2021; 98: 239–46.

22. Haaland RE, Chaowanachan T, Evans-Strickfaden T, et al. Carrageenan-based gel retains limited anti-HIV-1 activity 8-24 hours after vaginal application by HIV-infected Thai women enrolled in a phase I safety trial. J Acquir Immune Defic Syndr 2012; 61: e71–3.

23. Rodriguez A, Kleinbeck K, Mizenina O, et al. In vitro and in vivo evaluation of two carrageenan-based formulations to prevent HPV acquisition. Antiviral Res 2014; 108: 88–93.

24. Novetsky AP, Keller MJ, Gradissimo A, et al. In vitro inhibition of human papillomavirus following use of a carrageenan-containing vaginal gel. Gynecol Oncol 2016; 143: 313–8.

25. Kilmarx PH, van de Wijgert JHHM, Chaikummao S, et al. Safety and Acceptability of the Candidate Microbicide Carraguard in Thai Women. J Acquir Immune Defic Syndr 2006; 43: 327–34.

26. Laurie C, Franco E. The potential harms of personal lubricants. DST-J 2020; 32: 1–4.

27. Buck CB, Thompson CD, Roberts JN, et al. Carrageenan is a potent inhibitor of papillomavirus infection. PLoS Pathog 2006; 2: e69.

28. Perez-Andino J, Buck CB, Ribbeck K, Pérez-Andino J, Buck CB, Ribbeck K. Adsorption of human papillomavirus 16 to live human sperm. PLoS One 2009; 4: e5847.

29. Wang JW, Jagu S, Kwak K, et al. Preparation and properties of a papillomavirus infectious intermediate and its utility for neutralization studies. Virology 2014; 449: 304–16.

