## Supplementary material for "Self-applied carrageenan-based gel to prevent human papillomavirus infection in sexually active young women: Final analysis of efficacy and safety of a randomised controlled trial": CATCH Consort Checklist

### Supplementary Section 1

### Data collection and results using the daily calendar

The purpose of the CATCH calendar was to track participants' daily sexual activity, condom use, and compliance to the intervention. It was designed to be user friendly while conferring some degree of privacy through the use of picture icons (**Figure 1**). The calendar interface was set up to allow participants, when logging in securely through a computer or mobile device, to update their information. To minimize recall bias, participants were asked to update information for any given day within seven days, after which they were unable to view or modify that day's information. Thus, at any time, participants were able to enter or modify information for the seven previous days, i.e. participants needed to log in at least once a week.

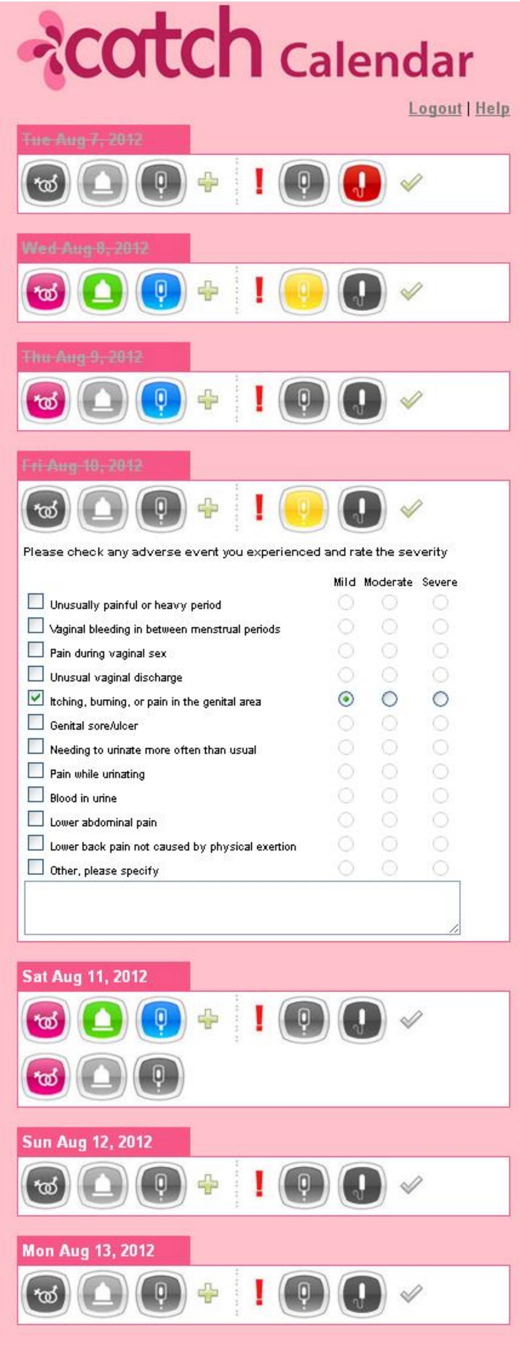


**Supplementary Figure 1. Interface of the CATCH daily calendar**

**<u>Supplementary Figure 1 Legend</u>**

Figure 1 shows an example of the daily calendar. Each icon has a mouse-over description. Using the top row as an example , the icons in sequence read:

"Press to indicate vaginal intercourse with a male partner on this day”

“Press if condoms were used during vaginal intercourse”

“Press if the CATCH gel was used before or during sexual activity”

“Press if you engaged in vaginal intercourse more than once in this day”

“Press to report adverse reactions to gel use”

“Press if the CATCH gel was used vaginally while not engaging in sexual activity” “Press if you were menstruating on this day”

“Press to cross out days that have passed” or if the. day is already crossed out “Press to edit this day’s information”.

The icons change from grey to colored when they are pressed. Strikethrough indicates the date is in the past.

If the participant pressed an icon to report adverse events (see example for August 10, 2012), the participant could then click a box to indicate which adverse event(s) they experienced, and also specify whether the adverse event was mild, moderate, or severe.

**Supplementary Table 1A. Number of entries in the daily calendar, by study arm.** The number of entries corresponds to the number of days participants filled in the calendar. The total number of follow-up days was 147,888.

| | Carrageenan | Placebo |
|---|---|---|
| Number of entries | 46,902 | 48,425 |

**Supplementary Table 1B. Adherence to daily calendar use, by study arm.** Adherence was calculated as the number of entries made between two consecutive study visits divided by the actual number of days between a study interval, multiplied by 100.

| Adherence to calendar use, n (%) | Carrageenan (Number of follow-up study visits=988) | Placebo (Number of follow-up study visits=1068) |
|---|---|---|
| <25 | 64 (6·5) | 73 (6·8) |
| 25 – 49·9 | 81 (8·2) | 91 (8·5) |
| 50 – 74·9 | 148 (15·0) | 164 (15·4) |
| 75 – 99.9 | 418 (42·3) | 483 (45·2) |
| 100 | 202 (20·5) | 172 (16·1) |
| No entries in interval | 75 (7·6) | 85 (8·0) |

**Supplementary Table 1C. Adherence to gel use, by study arm.** Adherence was calculated as the number of gel usages before/after vaginal intercourse divided by the number of vaginal intercourses within the study interval, multiplied by 100.

| Adherence to gel use, n (%) | Carrageenan (Number of follow-up study visits=988) | Placebo (Number of follow-up study visits=1068) |
|---|---|---|
| 0 | 141 (14·3) | 165 (15·5) |
| >0 – 24.9 | 43 (4·4) | 57 (5·3) |
| 25 – 49·9 | 88 (8·9) | 83 (7·8) |
| 50 – 74·9 | 112 (11·3) | 145 (13·6) |
| 75 – 99.9 | 103 (10·4) | 90 (8·4) |
| 100 | 187 (18·9) | 214 (20·0) |
| No intercourse in interval | 242 (24·5) | 231 (21·6) |
| No entries in interval | 72 (7·3) | 83 (7·8) |

### Supplementary Section 2

### Validation of HPV vaccination status

At the time of interim analysis, participants were emailed to confirm their HPV vaccination status. However, this procedure was later discontinued – starting September 25, 2019 – as the baseline and follow-up surveys were amended to include more detailed questions about HPV vaccination status, including the number of doses, brand name of the vaccine, and date of the first vaccination dose. There could be detection bias, as we did not email participants who reported not being vaccinated to verify their HPV vaccination status.

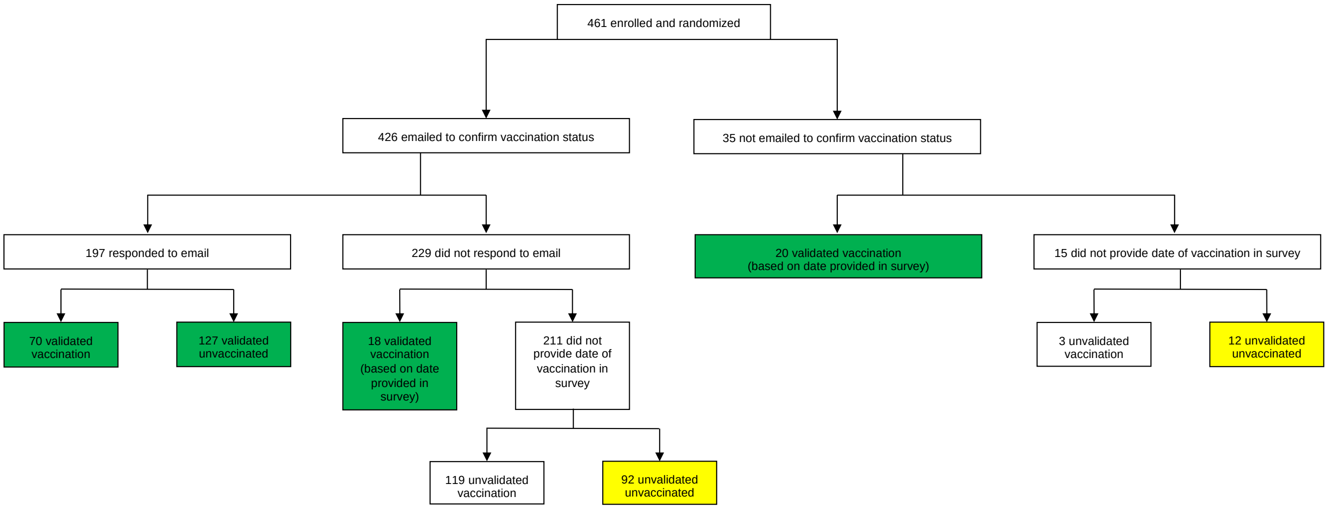


**Supplementary Figure 2**. Flowchart of participants through different methods of verifying HPV vaccination status

**<u>Supplementary Figure 2 Legend</u>**

Of the 426 participants emailed to confirm their vaccination status, 46·2% (197/426) responded to the email. Of the 229 who did not respond, 18 had provided the date of vaccination in the baseline or follow-up survey. Of the 35 participants who were not emailed, 57·1% (20/35) provided their date of vaccination in the baseline or follow-up survey. Boxes in green correspond to the most conservative validated definition of HPV vaccination status; these were included in the main Figure 1 and Table 3 of the manuscript. The combination of green and yellow boxes represent the liberal definition of HPV vaccination status.

**Supplementary Table 2A. HPV vaccination status reported in baseline survey or at screening, no validation**

| Vaccination status at baseline, n (%) | Carrageenan (n=227) | Placebo (n=234) |
|---|---|---|
| Vaccinated | 97 (42·7) | 120 (51·3) |
| Unvaccinated | 130 (57·3) | 114 (48·7) |

Imbalance but not significant ($p < 0.066$).

**Supplementary Table 2B. Comparison of HPV vaccination status at baseline to the validated vaccination status for participants who were emailed**

| | Validated status | |
|---|---|---|
| Vaccination status at baseline, n(%) | Vaccinated (n=70) | Unvaccinated (n=127) |
| Vaccinated | 56 | 4 |
| Unvaccinated | 14 | 123 |

We changed the HPV vaccination for 14 participants from unvaccinated to vaccinated and for 4 participants from vaccinated to unvaccinated.

**Supplementary Table 2C. Comparison of HPV vaccination using different definitions of validation from different sources**

| Validation description | HPV vaccination status at baseline | Carrageenan | Placebo |
|---|---|---|---|
| Validated by email | | n=97 | n=100 |
| | Vaccinated | 33 (34·0) | 37 (37·0) |
| | Unvaccinated | 64 (66·0) | 63 (63·0) |
| Validated by email or by providing dates of vaccination in survey(s) | | n=111 | n=124 |
| | Vaccinated | 47 (42·3) | 61 (49·2) |
| | Unvaccinated | 64 (57·7) | 63 (50·8) |
| Validated by email, by providing dates of vaccination in survey(s), or reporting they were unvaccinated in survey(s) | | n=174 | n=165 |
| | Vaccinated | 47 (27·0) | 61 (37·0) |
| | Unvaccinated | 127 (73·0) | 104 (63·0) |

**Supplementary Figure 3. Distribution of time between baseline and subsequent visits compared to the study schedule, by study arm**

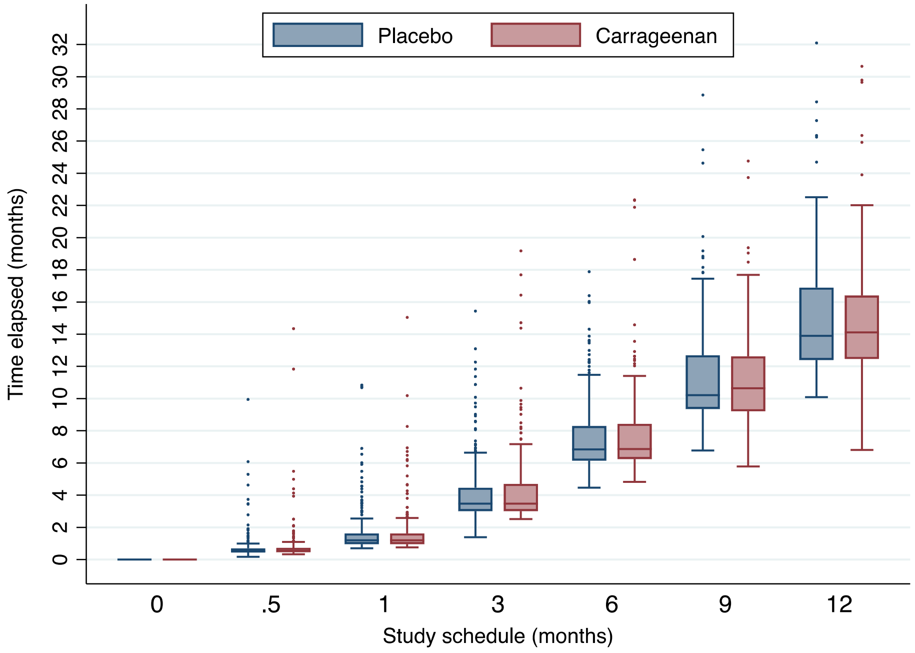


**Supplementary Figure 3 Legend**

The box plot shows on the x-axis the planned study schedule (baseline study visit and 6 follow-up visits at 0.5, 1, 3, 6, 9 and 12 months) relative to the actual time that elapsed between study visits on the y-axis. The midline of each box represents the median, the ends of the box represent the the first and third quartile, and the lower and upper whiskers range from the minimum to the maximum values observed (excluding outliers). The individual dots represent outliers.

**Supplementary Figure 4. Sub-group analyses of incidence/detection of HPV infections**

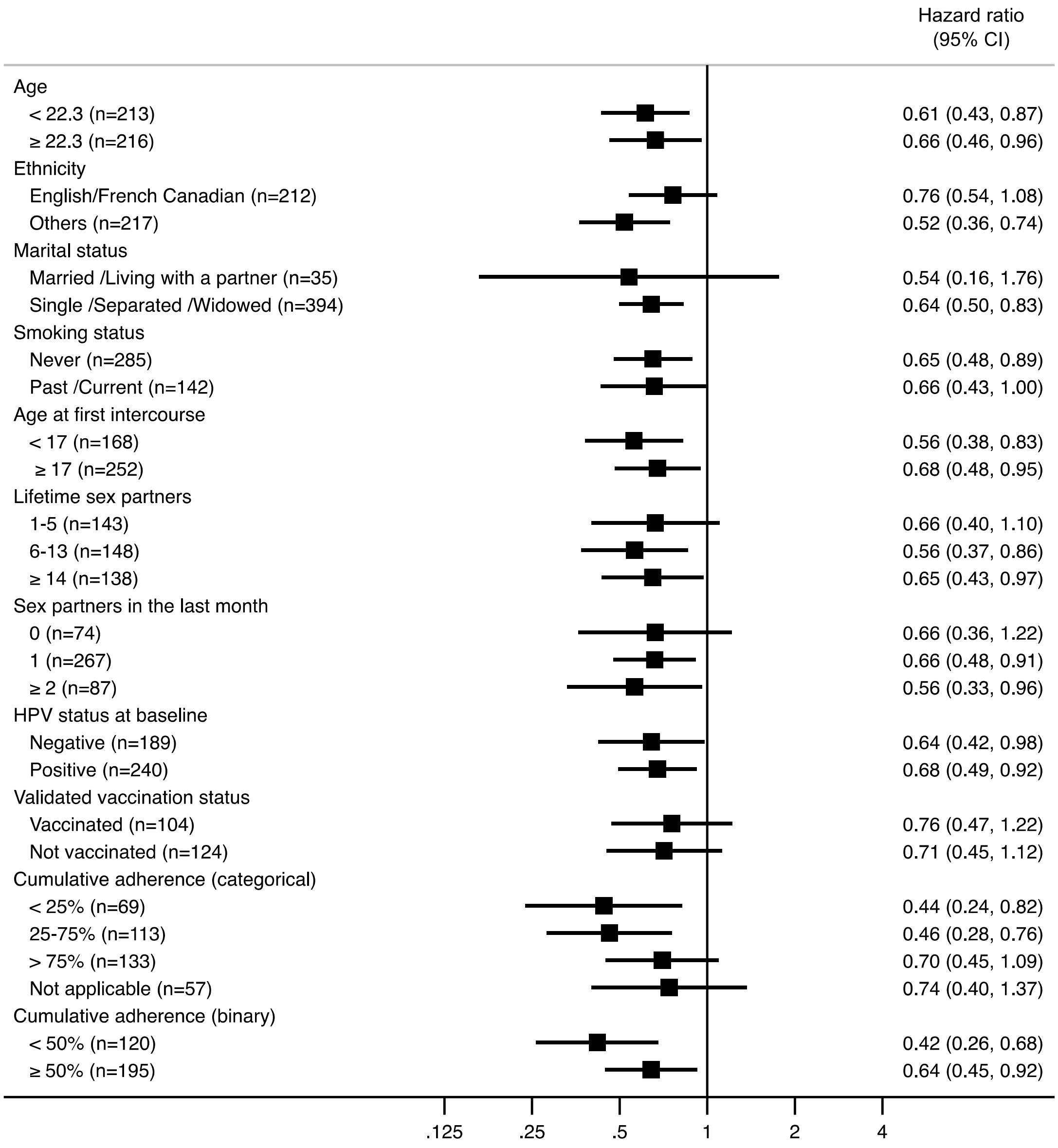


**<u>Supplementary Figure 4 Legend</u>**

The forest plot shows the results from sub-group analyses for the purpose of comparing the effect of carrageenan according to main baseline characteristics and cumulative adherence to follow-up based on calendar data.

**Supplementary Table 3A. HPV prevalence [n, (%)] by visit number in carrageenan arm**

| HPV type/groups | Visit 1 (n=224) | Visit 2 (n=202) | Visit 3 (n=187) | Visit 4 (n=170) | Visit 5 (n=151) | Visit 6 (n=140) | Visit 7 (n=126) |
|---|---|---|---|---|---|---|---|
| **HPV6** | 5 (2·2) | 4 (2·0) | 5 (2·7) | 5 (2·9) | 3 (2·0) | 3 (2·1) | 2 (1·6) |
| **HPV11** | 1 (0·5) | 1 (0·5) | 1 (0·5) | 2 (1·2) | 1 (0·7) | 0 (0) | 0 (0) |
| **HPV16** | 8 (3·6) | 11 (5·5) | 10 (5·4) | 12 (7·1) | 10 (6·6) | 8 (5·7) | 6 (4·8) |
| **HPV18** | 3 (1·3) | 2 (1·0) | 2 (1·1) | 1 (0·6) | 2 (1·3) | 1 (0·7) | 2 (1·6) |
| **HPV26** | 0 (0) | 0 (0) | 0 (0) | 1 (0·6) | 1 (0·7) | 1 (0·7) | 2 (1·6) |
| **HPV31** | 6 (2·7) | 3 (1·5) | 5 (2·7) | 6 (3·5) | 4 (2·7) | 4 (2·9) | 4 (3·2) |
| **HPV33** | 1 (0·5) | 0 (0) | 1 (0·5) | 1 (0·6) | 2 (1·3) | 3 (2·1) | 0 (0) |
| **HPV34** | 0 (0) | 0 (0) | 0 (0) | 0 (0) | 0 (0) | 0 (0) | 0 (0) |
| **HPV35** | 4 (1·8) | 0 (0) | 0 (0) | 1 (0·6) | 2 (1·3) | 1 (0·7) | 2 (1·6) |
| **HPV39** | 13 (5·8) | 10 (5·0) | 12 (6·4) | 13 (7·7) | 12 (8·0) | 10 (7·1) | 12 (9·5) |
| **HPV40** | 7 (3·1) | 6 (3·0) | 4 (2·1) | 3 (1·8) | 3 (2·0) | 5 (3·6) | 4 (3·2) |
| **HPV42** | 17 (7·6) | 13 (6·4) | 10 (5·4) | 12 (7·1) | 10 (6·6) | 9 (6·4) | 9 (7·1) |
| **HPV44** | 6 (2·7) | 4 (2·0) | 3 (1·6) | 3 (1·8) | 3 (2·0) | 3 (3·6) | 2 (1·6) |
| **HPV45** | 6 (2·7) | 3 (1·5) | 2 (1·1) | 4 (2·4) | 3 (2·0) | 5 (3·6) | 3 (2·4) |
| **HPV51** | 22 (9·8) | 16 (7·9) | 13 (7·0) | 12 (7·1) | 8 (5·3) | 10 (7·1) | 8 (6·4) |
| **HPV52** | 17 (7·6) | 12 (5·9) | 11 (5·9) | 10 (5·9) | 9 (6·0) | 7 (5·0) | 6 (4·8) |
| **HPV53** | 22 (9·8) | 18 (8·9) | 15 (8·0) | 13 (7·7) | 12 (8·0) | 12 (8·6) | 8 (6·4) |
| **HPV54** | 10 (4·5) | 8 (4·0) | 6 (3·2) | 9 (5·3) | 15 (9·9) | 10 (7·1) | 9 (7·1) |
| **HPV56** | 15 (6·7) | 16 (7·9) | 13 (7·0) | 10 (5·9) | 6 (4·0) | 6 (4·3) | 4 (3·2) |
| **HPV58** | 12 (5·4) | 9 (4·5) | 8 (4·3) | 6 (3·5) | 7 (4·6) | 4 (2·9) | 4 (3·2) |
| **HPV59** | 11 (4·9) | 12 (5·9) | 11 (5·9) | 10 (5·9) | 10 (6·6) | 11 (7·9) | 8 (6·4) |
| **HPV61** | 9 (4·0) | 7 (3·5) | 8 (4·3) | 8 (4·7) | 6 (4·0) | 4 (2·9) | 5 (4·0) |
| **HPV62** | 14 (6·3) | 11 (5·5) | 9 (4·8) | 6 (3·5) | 7 (4·6) | 5 (3·6) | 8 (6·4) |
| **HPV66** | 20 (8·9) | 17 (8·4) | 12 (6·4) | 12 (7·1) | 13 (8·6) | 8 (5·7) | 7 (5·6) |
| **HPV67** | 6 (2·7) | 6 (3·0) | 6 (3·2) | 11 (6·5) | 6 (4·0) | 4 (2·9) | 4 (3·2) |
| **HPV68** | 5 (2·2) | 5 (2·5) | 5 (2·7) | 3 (1·8) | 1 (0·7) | 1 (0·7) | 3 (2·4) |
| **HPV69** | 1 (0·5) | 2 (1·0) | 2 (1·1) | 1 (0·6) | 0 (0) | 1 (0·7) | 0 (0) |
| **HPV70** | 3 (1·3) | 4 (2·0) | 2 (1·1) | 0 (0) | 0 (0) | 0 (0) | 0 (0) |
| **HPV71** | 0 (0) | 0 (0) | 0 (0) | 0 (0) | 0 (0) | 1 (0·7) | 1 (0·8) |
| **HPV72** | 1 (0·5) | 1 (0·5) | 0 (0) | 0 (0) | 1 (0·7) | 1 (0·7) | 1 (0·8) |
| **HPV73** | 5 (2·2) | 7 (3·5) | 5 (2·7) | 7 (4·1) | 9 (6·0) | 4 (2·9) | 5 (4·0) |
| **HPV81** | 6 (2·7) | 2 (1·0) | 5 (2·7) | 3 (1·8) | 3 (2·0) | 3 (2·1) | 4 (3·2) |
| **HPV82** | 4 (1·8) | 0 (0) | 2 (1·1) | 2 (1·2) | 3 (2·0) | 6 (4·3) | 5 (4·0) |
| **HPV83** | 3 (1·3) | 3 (1·5) | 3 (1·6) | 4 (2·4) | 6 (4·0) | 5 (3·6) | 3 (2·4) |
| **HPV84** | 20 (8·9) | 21 (10·4) | 19 (10·2) | 20 (11·8) | 13 (8·6) | 13 (9·3) | 6 (4·8) |
| **HPV89** | 22 (9·8) | 22 (10·9) | 17 (9·1) | 19 (11·2) | 14 (9·3) | 9 (6·4) | 9 (7·1) |
| **Negative** | 109 (48·7) | 100 (49·5) | 92 (49·2) | 76 (44·7) | 70 (46·4) | 69 (49·3) | 61 (48·4) |
| **Any HPV** | **115 (50·5)** | **102 (50·5)** | **95 (50·8)** | **94 (55·3)** | **81 (53·6)** | **71 (50·7)** | **65 (51·6)** |
| **Subgenus 1[a]** | 38 (17·0) | 31 (15·4) | 27 (14·4) | 30 (17·7) | 30 (19·9) | 26 (18·6) | 19 (15·1) |
| **Subgenus 2[b]** | 96 (42·9) | 81 (40·1) | 75 (40·1) | 73 (42·9) | 61 (40·4) | 60 (42·9) | 53 (42·1) |
| **Subgenus 3[c]** | 60 (26·8) | 52 (25·7) | 46 (24·6) | 47 (27·7) | 37 (24·5) | 30 (21·4) | 31 (24·6) |
| **Missing[d]** | 3 (1·3) | 6 (2·9) | 3 (1·6) | 1 (0·6) | 1 (0·7) | 0 (0) | 1 (0·8) |

[a] Subgenus 1 group consists of HPVs 6, 11, 40, 42, 44, and 54.
[b] Subgenus 2 group consists of HPVs 16, 18, 26, 31, 33, 34, 35, 39, 45, 51, 52, 53, 56, 58, 59, 66, 67, 68, 69, 70, 73, and 82.
[c] Subgenus 3 group consists of HPVs 61, 62, 71, 72, 81, 83, 84, and 89.
[d] Missing results correspond to invalid or mishandled samples. Missing samples were not included in percentages.

**Supplementary Table 3B. HPV prevalence [n, (%)] by visit number in placebo arm**

| HPV type/groups | Visit 1 (n=234) | Visit 2 (n=221) | Visit 3 (n=208) | Visit 4 (n=190) | Visit 5 (n=164) | Visit 6 (n=151) | Visit 7 (n=130) |
|---|---|---|---|---|---|---|---|
| **HPV6** | 5 (2·1) | 5 (2·3) | 5 (2·4) | 5 (2·6) | 6 (3·7) | 6 (4·0) | 5 (3·9) |
| **HPV11** | 0 (0) | 0 (0) | 0 (0) | 1 (0·5) | 1 (0·6) | 1 (0·7) | 1 (0·8) |
| **HPV16** | 10 (4·3) | 12 (5·4) | 12 (5·8) | 10 (5·3) | 11 (6·7) | 10 (6·6) | 7 (5·4) |
| **HPV18** | 6 (2·6) | 3 (1·4) | 4 (1·9) | 4 (2·1) | 2 (1·2) | 4 (2·7) | 2 (1·5) |
| **HPV26** | 1 (0·4) | 1 (0·5) | 0 (0) | 1 (0·5) | 1 (0·6) | 0 (0) | 0 (0) |
| **HPV31** | 7 (3·0) | 6 (2·7) | 10 (4·8) | 11 (5·8) | 9 (5·5) | 10 (6·6) | 1 (0·8) |
| **HPV33** | 2 (0·9) | 4 (1·8) | 3 (1·4) | 4 (2·1) | 3 (1·8) | 1 (0·7) | 0 (0) |
| **HPV34** | 1 (0·4) | 1 (0·5) | 1 (0·5) | 1 (0·5) | 0 (0) | 1 (0·7) | 3 (2·3) |
| **HPV35** | 7 (3·0) | 6 (2·7) | 6 (2·9) | 4 (2·1) | 3 (1·8) | 3 (2·0) | 3 (2·3) |
| **HPV39** | 16 (6·8) | 15 (6·8) | 16 (7·7) | 9 (4·7) | 12 (7·3) | 11 (7·3) | 10 (7·7) |
| **HPV40** | 8 (3·4) | 7 (3·2) | 10 (4·8) | 11 (5·8) | 6 (3·7) | 7 (4·6) | 3 (2·3) |
| **HPV42** | 23 (9·8) | 26 (11·8) | 20 (9·6) | 17 (9·0) | 16 (9·8) | 18 (11·9) | 17 (13·1) |
| **HPV44** | 8 (3·4) | 13 (5·9) | 10 (4·8) | 11 (5·8) | 5 (3·1) | 7 (4·6) | 3 (2·3) |
| **HPV45** | 6 (2·6) | 5 (2·3) | 5 (2·4) | 6 (3·2) | 7 (4·3) | 6 (4·0) | 2 (1·5) |
| **HPV51** | 34 (14·5) | 30 (13·6) | 25 (12·0) | 23 (12·1) | 16 (9·8) | 14 (9·3) | 8 (6·2) |
| **HPV52** | 13 (5·6) | 16 (7·2) | 11 (5·3) | 11 (5·8) | 9 (5·5) | 9 (6·0) | 11 (8·5) |
| **HPV53** | 31 (13·3) | 32 (14·5) | 25 (12·0) | 28 (14·7) | 28 (17·1) | 18 (11·9) | 15 (11·5) |
| **HPV54** | 18 (7·7) | 15 (6·8) | 16 (7·7) | 18 (9·5) | 16 (9·8) | 12 (8·0) | 14 (10·8) |
| **HPV56** | 6 (2·6) | 6 (2·7) | 6 (2·9) | 7 (3·7) | 3 (1·8) | 6 (4·0) | 3 (2·3) |
| **HPV58** | 13 (5·6) | 13 (5·9) | 10 (4·8) | 10 (5·3) | 12 (7·3) | 11 (7·3) | 9 (6·9) |
| **HPV59** | 19 (8·1) | 18 (8·1) | 15 (7·2) | 12 (6·3) | 14 (8·5) | 8 (5·3) | 5 (3·9) |
| **HPV61** | 10 (4·3) | 10 (4·5) | 12 (5·8) | 15 (7·9) | 12 (7·3) | 9 (6·0) | 6 (4·6) |
| **HPV62** | 21 (9·0) | 19 (8·6) | 12 (5·8) | 21 (11·1) | 18 (11·0) | 15 (9·9) | 13 (10·0) |
| **HPV66** | 23 (9·8) | 27 (12·2) | 22 (10·6) | 18 (9·5) | 11 (6·7) | 12 (8·0) | 12 (9·2) |
| **HPV67** | 13 (5·6) | 13 (5·9) | 15 (7·2) | 11 (5·8) | 8 (4·9) | 4 (2·7) | 5 (3·9) |
| **HPV68** | 6 (2·6) | 7 (3·2) | 4 (1·9) | 3 (1·6) | 4 (2·4) | 3 (2·0) | 2 (1·5) |
| **HPV69** | 0 (0) | 0 (0) | 0 (0) | 0 (0) | 0 (0) | 0 (0) | 1 (0·8) |
| **HPV70** | 3 (1·3) | 4 (1·8) | 5 (2·4) | 2 (1·1) | 3 (1·8) | 3 (2·0) | 1 (0·8) |
| **HPV71** | 0 (0) | 0 (0) | 0 (0) | 0 (0) | 0 (0) | 1 (0·7) | 1 (0·8) |
| **HPV72** | 0 (0) | 1 (0·5) | 0 (0) | 0 (0) | 0 (0) | 0 (0) | 1 (0·8) |
| **HPV73** | 14 (6·0) | 14 (6·3) | 12 (5·8) | 9 (4·7) | 10 (6·1) | 12 (8·0) | 6 (4·6) |
| **HPV81** | 8 (3·4) | 7 (3·2) | 6 (2·9) | 6 (3·2) | 5 (3·1) | 2 (1·3) | 6 (4·6) |
| **HPV82** | 7 (3·0) | 6 (2·7) | 11 (5·3) | 9 (4·7) | 7 (4·3) | 8 (5·3) | 1 (0·8) |
| **HPV83** | 6 (2·6) | 5 (2·3) | 6 (2·9) | 5 (2·6) | 5 (3·1) | 5 (3·3) | 6 (4·6) |
| **HPV84** | 29 (12·4) | 25 (11·3) | 25 (12·0) | 21 (11·1) | 18 (11·0) | 17 (11·3) | 14 (10·8) |
| **HPV89** | 27 (11·5) | 37 (16·7) | 33 (15·9) | 21 (11·1) | 23 (1·40) | 19 (12·6) | 11 (8·5) |
| **Negative** | 93 (39·7) | 78 (35·3) | 73 (35·1) | 64 (33·7) | 57 (34·8) | 60 (39·7) | 56 (43·1) |
| **Any HPV** | **141 (60·3)** | **143 (64·7)** | **135 (64·9)** | **126 (66·3)** | **107 (65·2)** | **91 (60·3)** | **74 (56·9)** |
| **Subgenus 1[a]** | 55 (23·5) | 55 (24·9) | 52 (25·0) | 49 (25·8) | 44 (26·8) | 40 (26·8) | 33 (25·4) |
| **Subgenus 2[b]** | 114 (48·7) | 117 (52·9) | 110 (52·9) | 103 (54·2) | 89 (54·3) | 74 (49·0) | 55 (42·3) |
| **Subgenus 3[c]** | 76 (32·5) | 78 (35·3) | 71 (34·1) | 63 (33·2) | 58 (35·4) | 52 (34·4) | 46 (35·4) |
| **Missing[d]** | 0 (0) | 0 (0) | 1 (0·5) | 1 (0·5) | 2 (1·2) | 0 (0) | 0 (0) |

[a] Subgenus 1 group consists of HPVs 6, 11, 40, 42, 44, and 54.
[b] Subgenus 2 group consists of HPVs 16, 18, 26, 31, 33, 34, 35, 39, 45, 51, 52, 53, 56, 58, 59, 66, 67, 68, 69, 70, 73, and 82.
[c] Subgenus 3 group consists of HPVs 61, 62, 71, 72, 81, 83, 84, and 89.
[d] Missing results correspond to invalid or mishandled samples. Missing samples were not included in percentages.

**Supplementary Table 4. Characteristics of participants at follow-up, overall and by study arm**

| | Overall (n=461) | Carrageenan (n=227) | Placebo (n=234) |
|---|---|---|---|
| **Follow-up time, months** | | | |
| Mean (SD) | 10·6 (6·8) | 10·4 (7·0) | 10·8 (6·6) |
| Median (IQR) | 12·2 (11·1) | 12·2 (11·8) | 12·2 (10·2) |
| **Number of visits/participant** | | | |
| Mean (SD) | 5·5 (2·1) | 5·4 (2·2) | 5·6 (1·9) |
| Median (IQR) | 7 (3) | 7 (3) | 7 (3) |
| **Mean time between visits (SD), weeks** | | | |
| Between visit 1 and 2 | 3·6 (5·2) | 3·8 (6·1) | 3·4 (4·1) |
| Between visit 2 and 3 | 4·1 (5·0) | 4·1 (4·9) | 4·1 (5·0) |
| Between visit 3 and 4 | 11·6 (7·5) | 11·9 (7·9) | 11·4 (7·2) |
| Between visit 4 and 5 | 15·2 (6·5) | 15·3 (6·4) | 15·2 (6·6) |
| Between visit 5 and 6 | 16·0 (8·2) | 15·9 (7·3) | 16·1 (9·1) |
| Between visit 6 and 7 | 16·5 (8·7) | 16·6 (8·6) | 16·4 (8·7) |
| **Ever anal intercourse over follow-up[a]** | **(n=428)** | **(n=207)** | **(n=221)** |
| Yes | 124 (29·0) | 54 (26·1) | 70 (31·7) |
| No | 304 (71·0) | 153 (73·9) | 151 (68·3) |
| **Adherence[b] in the 7 days preceding each visit, n (%)** | **(n=2056)** | **(n=988)** | **(n=1068)** |
| >75% | 544 (26·5) | 257 (26·0) | 287 (26·9) |
| 25-75% | 258 (12·6) | 118 (11·9) | 140 (13·1) |
| <25% | 325 (15·8) | 143 (14·5) | 182 (17·0) |
| Did not have intercourse | 915 (44·5) | 459 (46·5) | 456 (42·7) |
| Not reported | 14 (0·7)_ | 11 (1·11) | 3 (0·3) |

[a] Data not available for 32 participants (19 in the carrageenan and 13 in the placebo arm) due to no follow-up visits, and for 1 participant due to non-response.
[b] Adherence was defined as the number of times the gel was used before or during vaginal intercourse divided by the number of vaginal intercourses in the 7 days prior to the study visit. Observations where participants did not provide data on gel use and/or intercourse were classified as not reported. This data was obtained from the follow-up surveys.
IQR: interquartile range.

**Supplementary Table 5. Adverse events reported in the calendar, overall and by grade, shown as number of adverse events reported (number of participants affected)**

| | Mild | Moderate | Severe | Not Graded | Total |
|---|---|---|---|---|---|
| **Carrageenan group** | | | | | |
| Any | 89 (21) | 40 (19) | 24 (9) | 75 (22) | 228 (37) |
| Unusually heavy or painful period | 2 (1) | 1 (1) | 0 (0) | 0 (0) | 3 (1) |
| Vaginal bleeding in between menstrual periods | 16 (5) | 0 (0) | 0 (0) | 1 (1) | 17 (6) |
| Pain during vaginal sex | 8 (6) | 5 (4) | 4 (3) | 5 (3) | 22 (13) |
| Unusual vaginal discharge | 16 (5) | 6 (6) | 2 (2) | 8 (2) | 32 (11) |
| Itching, burning, or pain in the genital area | 35 (16) | 18 (9) | 14 (8) | 11 (5) | 78 (23) |
| Genital sore/ulcer | 1 (1) | 1 (1) | 0 (0) | 0 (0) | 2 (2) |
| Needing to urinate more often than usual | 2 (2) | 2 (2) | 0 (0) | 0 (0) | 4 (3) |
| Pain while urinating | 3 (3) | 0 (0) | 2 (2) | 5 (1) | 10 (6) |
| Blood in urine | 1 (1) | 0 (0) | 1 (1) | 3 (1) | 5 (3) |
| Lower abdominal pain | 5 (4) | 5 (3) | 1 (1) | 0 (0) | 11 (5) |
| Lower back pain not caused by physical exertion | 0 (0) | 0 (0) | 0 (0) | 0 (0) | 0 (0) |
| Other | 0 (0) | 2 (2) | 0 (0) | 42 (20) | 44 (21) |
| **Placebo group** | | | | | |
| Any | 48 (21) | 57 (22) | 27 (12) | 56 (23) | 188 (39) |
| Unusually heavy or painful period | 4 (4) | 9 (3) | 1 (1) | 1 (1) | 15 (8) |
| Vaginal bleeding in between menstrual periods | 9 (5) | 2 (2) | 1 (1) | 1 (1) | 13 (8) |
| Pain during vaginal sex | 0 (0) | 3 (3) | 6 (3) | 1 (1) | 10 (6) |
| Unusual vaginal discharge | 9 (2) | 9 (4) | 3 (2) | 0 (0) | 21 (4) |
| Itching, burning, or pain in the genital area | 17 (11) | 19 (13) | 12 (6) | 5 (5) | 53 (19) |
| Genital sore/ulcer | 3 (3) | 4 (1) | 0 (0) | 0 (0) | 7 (3) |
| Needing to urinate more often than usual | 2 (2) | 2 (2) | 1 (1) | 5 (3) | 10 (7) |
| Pain while urinating | 0 (0) | 2 (2) | 1 (1) | 3 (1) | 6 (3) |
| Blood in urine | 0 (0) | 0 (0) | 0 (0) | 1 (1) | 1 (1) |
| Lower abdominal pain | 3 (3) | 2 (2) | 0 (0) | 2 (2) | 7 (3) |
| Lower back pain not caused by physical exertion | 0 (0) | 2 (1) | 0 (0) | 1 (1) | 3 (2) |
| Other | 1 (1) | 3 (3) | 2 (2) | 36 (18) | 42 (21) |

**Supplementary Table 6. Participant withdrawal overall and stratified by reporting of adverse events and reporting of difficulties with gel use, by study arm**

| | Total (n=461) | Carrageenan (n=227) | Placebo (n=234) |
|---|---|---|---|
| Overall | | | |
| Completed | 257 (55·8) | 127 (56·0) | 130 (55·6) |
| Withdrew | 204 (44·3) | 100 (44·1) | 104 (44·4) |
| Reported adverse events | | | |
| Completed | 107 (62·2) | 50 (63·3) | 57 (61·3) |
| Withdrew | 65 (37·8) | 29 (36·7) | 36 (38·7) |
| Did not report adverse events | | | |
| Completed | 150 (51·9) | 77 (52·0) | 73 (51·8) |
| Withdrew | 139 (48·1) | 71 (48·0) | 68 (48·2) |
| Reported difficulty using gel before, during or after intercourse[a] | | | |
| Completed | 209 (62·4) | 101 (63·5) | 108 (61·4) |
| Withdrew | 126 (37·6) | 58 (36·5) | 68 (38·6) |
| Did not report difficulty using gel before, during or after intercourse[a] | | | |
| Completed | 48 (53.9) | 26 (55·3) | 22 (52·4) |
| Withdrew | 41 (46·1) | 21 (44·7) | 20 (47·6) |

[a] Because reporting difficulty using gel was a follow-up survey question, only participants with a follow-up visit were included in the denominator: excluded 32 participants (19 in the carrageenan and 13 in the placebo arm). An additional 5 participants (2 in the carrageenan and 3 in the placebo) were excluded due to non-response.

**Supplementary Table 7. Reported difficulty using the study gel before, during, or after intercourse [n (%)] at the person- and study-visit level, by study arm**

| | Person-level[a] | | Study visit-level[b] | |
|---|---|---|---|---|
| | Carrageenan (n=206) | Placebo (n=218) | Carrageenan (n=871) | Placebo (n=962) |
| **Ever reported difficulty with gel** | 159 (77·2) | 176 (80·7) | NA | NA |
| Application of the CATCH gel is too difficult | 17 (8·3) | 22 (10·1) | 28 (3·2) | 33 (3·4) |
| The packaging is too inconvenient | 19 (9·2) | 19 (8·7) | 37 (4·3) | 32 (3·3) |
| You did not have the CATCH gel on you at the time of intercourse | 98 (47·6) | 117 (53·7) | 231 (26·5) | 282 (29·3) |
| You forgot to use the CATCH gel | 95 (46·1) | 111 (50·9) | 212 (24·3) | 248 (25·8) |
| You did not want to use lubricants | 54 (26·2) | 85 (39·0) | 105 (12·1) | 184 (19·1) |
| You preferred other brands to the CATCH gel | 24 (11·7) | 31 (14·2) | 44 (5·1) | 60 (6·2) |
| You think that the quality of the CATCH gel is poor (e.g., odour, feel, etc.) | 35 (17·0) | 66 (30·3) | 66 (7·6) | 150 (15·6) |
| Use of the gel caused discomfort/adverse reactions to you | 22 (10·7) | 27 (12·4) | 24 (2·8) | 35 (3·6) |
| Partner(s) did not want to use lubricants | 36 (17·5) | 54 (24·8) | 67 (7·7) | 93 (9·7) |
| Partner(s) is/are allergic to ingredients of the CATCH gel | 0 (0) | 1 (0·5) | 0 (0) | 1 (0·1) |
| Partner(s) preferred other brands to the CATCH gel | 18 (8·7) | 16 (7·3) | 34 (3·9) | 22 (2·3) |
| Partner(s) think(s) that the quality of the CATCH gel is poor (e.g., odour, feel, etc.) | 22 (10·7) | 31 (14·2) | 37 (4·3) | 54 (5·6) |
| Use of the gel caused discomfort/adverse reactions to your partner(s) | 5 (2·4) | 5 (2·3) | 5 (0·6) | 6 (0·6) |
| Other | 27 (13·1) | 21 (9·6) | 38 (4·4) | 26 (2·7) |

[a] Participants excluded: 19 participants in the carrageenan arm and 13 in the placebo arm only had 1 visit, and 2 participants in the carrageenan and 3 participants in the placebo never responded to this question.
[b] There were 117 study visits in the carrageenan and 106 in the placebo did not respond.
NA: not applicable.

### Supplementary Table 8. Adherence to gel use before/during intercourse overall and cumulative adherence prior to failure or censoring

| Source | Adherence definition | | Categorization | Number of participants | Adherence (%) | Hazard ratio (95% CI) |
|---|---|---|---|---|---|---|
| Follow-up survey | number of gel uses / number of vaginal intercourses reported **one week prior to study visit** | overall[a] | NA | 191 | >50 | 0.69 (0.47-1.02) |
| | | cumulative[b] | categorical | 65 | <25 | 0.57 (0.30-1.07) |
| | | | | 103 | 25-75 | 0.52 (0.29-0.94) |
| | | | | 155 | >75 | 0.72 (0.47-1.08) |
| | | | | 98 | NA | 0.64 (0.40-1.04) |
| | | | | 8 | missing data | NR |
| | | | binary | 112 | <50 | 0.58 (0.35-0.98) |
| | | | | 211 | ≥50 | 0.64 (0.44-0.92) |
| Calendar | cumulative number of gel uses / number of vaginal intercourses **between two consecutive study visits** | overall[a] | NA | 183 | >50 | 0.60 (0.41-0.89) |
| | | cumulative[b] | categorical | 69 | <25 | 0.44 (0.24-0.82) |
| | | | | 113 | 25-75 | 0.46 (0.28-0.76) |
| | | | | 133 | >75 | 0.70 (0.45-1.10) |
| | | | | 57 | NA | 0.74 (0.40-1.37) |
| | | | | 57 | missing data | 1.14 (0.47-2.76) |
| | | | binary | 120 | <50 | 0.42 (0.26-0.68) |
| | | | | 195 | ≥50 | 0.64 (0.45-0.92) |

Data on adherence was collected in both the follow-up surveys and daily calendar.

[a] Overall adherence to gel use before/during intercourse over all study visits. These analyses were restricted to participants who were >50% adherent over all study visits.

[b] Cumulative adherence to gel use before/during intercourse prior to failure or censoring.

Not applicable if participant did not report intercourse prior to acquiring an incident infection or censoring.

Not reported, as there were only 8 participants in this category, so Cox model was not run.

NA: not applicable, NR: not reported.

### Supplementary Table 9. References of studies that reported on carrageenan's anti-HPV activity

| Study type | Reference |
|---|---|
| *in vitro* | 1. Buck CB, Thompson CD, Roberts JN, et al. Carrageenan is a potent inhibitor of papillomavirus infection. PLoS Pathog 2006; **2**: e69.<br>2. Perez-Andino J, Buck CB, Ribbeck K, Pérez-Andino J, Buck CB, Ribbeck K. Adsorption of human papillomavirus 16 to live human sperm. PLoS One 2009; **4**: e5847.<br>3. Cruz L, Meyers C. Differential dependence on host cell glycosaminoglycans for infection of epithelial cells by high-risk HPV types. PLoS One 2013; **8**: e68379.<br>4. Kwak K, Jiang R, Wang JW, Jagu S, Kirnbauer R, Roden RBS. Impact of inhibitors and L2 antibodies upon the infectivity of diverse alpha and beta human papillomavirus types. PLoS One 2014; **9**: e97232.<br>5. Lal M, Lai M, Ugaonkar S, et al. Development of a Vaginal Fast-Dissolving Insert Combining Griffithsin and Carrageenan for Potential Use Against Sexually Transmitted Infections. J Pharm Sci 2018; **107**: 2601–10.<br>6. Wang S, Lu Z, Wang S, et al. The inhibitory effects and mechanisms of polymannuroguluronate sulfate against human papillomavirus infection in vitro and in vivo. Carbohydr Polym 2020; **241**: 116365. |
| *in vitro* and *ex vivo* | 1. Ugaonkar SR, Wesenberg A, Wilk J, et al. A novel intravaginal ring to prevent HIV-1, HSV-2, HPV, and unintended pregnancy. J Control Release; 2015. p. 57–68.<br>2. Novetsky AP, Keller MJ, Gradissimo A, et al. In vitro inhibition of human papillomavirus following use of a carrageenan-containing vaginal gel. Gynecol Oncol 2016; **143**: 313–8.<br>3. Friedland BA, Hoesley CJ, Plagianos M, et al. First-in-Human Trial of MIV-150 and Zinc Acetate Coformulated in a Carrageenan Gel. J Acquir Immune Defic Syndr 2016; **73**: 489–96.<br>4. Teleshova N, Keller MJ, Romero JAF, et al. Results of a phase 1, randomized, placebocontrolled first-in-human trial of griffithsin formulated in a carrageenan vaginal gel. PLoS One; 2022; **17**: e0261775. |
| *in vitro* and *in vivo* | 1. Rodriguez A, Kleinbeck K, Mizenina O, et al. In vitro and in vivo evaluation of two carrageenan-based formulations to prevent HPV acquisition. Antiviral Res 2014; **108**: 88–93.<br>2. Wang JW, Jagu S, Kwak K, et al. Preparation and properties of a papillomavirus infectious intermediate and its utility for neutralization studies. Virology 2014; **449**: 304–16.<br>3. Kizima L, Rodríguez A, Kenney J, et al. A potent combination microbicide that targets SHIV-RT, HSV-2 and HPV. PLoS One 2014; **9**: e94547.<br>4. Levendosky K, Mizenina O, Martinelli E, et al. Griffithsin and Carrageenan Combination To Target Herpes Simplex Virus 2 and Human Papillomavirus. Antimicrob Agents Chemother 2015; **59**: 7290–8.<br>5. Kines RC, Cerio RJ, Roberts JN, et al. Human papillomavirus capsids preferentially bind and infect tumor cells. Int J Cancer 2016; **138**: 901–11. |
| *in vivo* | 1. Roberts JN, Buck CB, Thompson CD, et al. Genital transmission of HPV in a mouse model is potentiated by nonoxynol-9 and inhibited by carrageenan. Nat Med 2007; **7**: 857–61.<br>2. Roberts JN, Kines RC, Katki HA, Lowy DR, Schiller JT. Effect of Pap Smear Collection and Carrageenan on Cervicovaginal Human Papillomavirus-16 Infection in a Rhesus Macaque Model. J Natl Cancer Inst 2011; **103**: 737–43.<br>3. Derby N, Lal M, Aravantinou M, et al. Griffithsin carrageenan fast dissolving inserts prevent SHIV HSV-2 and HPV infections in vivo. Nature Commun 2018; **9**: 3881. |
| clinical | 1. Marais D, Gawarecki D, Allan B, et al. The effectiveness of Carraguard, a vaginal microbicide, in protecting women against high-risk human papillomavirus infection. Antivir Ther 2011; **16**: 1219–26.<br>2. Perino A, Consiglio P, Maranto M, et al. Impact of a new carrageenan-based vaginal microbicide in a female population with genital HPV-infection: first experimental results. Eur Rev Med Pharmacol Sci 2019; **23**: 6744–52.<br>3. Magnan S, Tota JE, El-Zein M, et al. Efficacy of a Carrageenan gel Against Transmission of Cervical HPV (CATCH): interim analysis of a randomized, double-blind, placebo-controlled, phase 2B trial. Clin Microbiol Infect 2019; **25**: 210–6.<br>4. Laurie C, El-Zein M, Tota JE, et al. Efficacy of a carrageenan gel in preventing anal human papillomavirus (HPV) infection: interim analysis of the Lubricant Investigation in Men to Inhibit Transmission of HPV Infection (LIMIT-HPV) randomised controlled trial. Sex Transm Infect 2021; **98**: 239–246.<br>5. Laurie C, El-Zein M, Tota J, et al. Efficacy of a carrageenan gel in increasing clearance of anal HPV infections in men: interim analysis of a double-blind randomized controlled trial. J Infect Dis 2022; **227**: 402–406. |
